# THR-6E: A Six-Gene Cell-of-Origin Signature Stratifies Risk and Predicts Systemic Therapy Response in ER+/HER2− Breast Cancer

**DOI:** 10.64898/2026.01.31.26345244

**Authors:** Priyanka Vasanthakumari, Itzel Valencia, Mohamed Omar, Tan A. Ince

## Abstract

**Background:** Genomic assays such as Oncotype DX, MammaPrint, and Prosigna have transformed risk stratification and treatment selection in early-stage, estrogen receptor-positive (ER+), HER2-negative breast cancers by enabling more precise prognostication and chemotherapy de-escalation in selected patients. However, their clinical utility is limited in lymph nodes positive disease. A major unmet need is the development of compact, mechanistically grounded biomarkers that extend risk and treatment-response prediction to clinically challenging ER+/HER2− subgroups, including lymph node–positive patients.

**Methods:** Building on a cell-of-origin framework, we previously established a 70-gene triple hormone receptor (THR; ER, AR, VDR) signature (THR-70) that reflects luminal epithelial differentiation programs and is prognostic across breast cancer subtypes. Here, we refined this framework using interactome-guided clustering to derive a six-gene cell-of-origin signature (THR-6E: KIF4A, KIF2C, CDC20, FAM64A, TPX2, and LMNB2). We evaluated the prognostic and predictive performance of THR-6E across >7,000 breast cancer cases from multiple independent cohorts, assessed treatment-response prediction using endocrine- and chemotherapy-annotated datasets, and performed independent validation in the I-SPY2 adaptive clinical trial.

**Findings:** THR-6E robustly stratifies relapse-free survival (RFS) in ER+/HER2− breast cancer independent of tumor grade, proliferation status, and subtype. Hazard ratios for RFS were 2.41 (p<1×10⁻¹⁶), 1.61 (p=4.9×10⁻⁴), and 1.50 (p=6.2×10⁻³) for grades 1, 2, and 3, respectively, and 2.16 and 1.33 for Luminal A and Luminal B subtypes. THR-6E maintained predictive value across endocrine- and chemotherapy-treated, untreated, lymph node-positive, and lymph node-negative subgroups. Beyond prognosis, THR-6E predicted endocrine therapy response in ER+/HER2−, node-negative disease and chemotherapy response in ER+/HER2−, node-positive disease, achieving approximately 70% sensitivity and specificity (AUC=0.714, p=3.6×10⁻⁶), with consistent performance across taxane-, anthracycline-, and FEC-based regimens (AUCs 0.71–0.72). Single-cell transcriptomic and proteomic analyses demonstrated that THR-6E expression is specific to normal and malignant breast glandular epithelium, preserved during transformation, and further enriched in metastatic disease. Consistent with a cell-of-origin program, THR-6E genes were rarely mutated in breast cancer and retained normal tissue-like co-expression patterns. In the I-SPY2 trial, THR-6E achieved robust prediction of pathologic complete response across multiple treatment arms (AUCs 0.72–0.94), with an overall AUC of 0.741.

**Interpretation:** These results support a cell-of-origin–anchored approach to biomarker development and challenge purely tissue-agnostic models of therapeutic response. THR-6E represents a compact, biologically interpretable signature that extends prognostic and predictive assessment to clinically relevant ER+/HER2− subgroups, including lymph node–positive disease. Its mechanistic grounding and stable performance across cohorts position THR-6E as a complementary tool to existing multigene assays, warranting prospective diagnostic accuracy studies to define its clinical utility and impact on treatment decision-making.

## BACKGROUND

Advances in molecular profiling have reshaped risk assessment and treatment selection in breast cancer, moving beyond clinicopathologic factors to incorporate gene expression-based tools that refine recurrence risk and inform escalation or deescalation of systemic therapy. In early-stage, ER-positive (ER+)/HER2-negative (HER2−) disease, the Oncotype DX’s Recurrence Score (RS) was developed to predict distant recurrence risk and chemotherapy benefit (1–3) and was prospectively tested in the landmark TAILORx trial, which established that nearly 70% of women with early-stage, ER+/HER2− breast cancer can safely omit chemotherapy without compromising outcomes (4). Similarly, MammaPrint provided clinically actionable prognostic stratification and was evaluated in the MINDACT trial, showing that patients classified as clinically high-risk but genomically low-risk can safely avoid chemotherapy (5). Prosigna (PAM-50) has also been validated across settings to generate a risk of recurrence (ROR) score that captures both early and late recurrence risk in ER+/HER2− breast cancer (6). Collectively, these assays have changed practice by altering adjuvant treatment recommendations in a meaningful fraction of tested patients, most often by reducing chemotherapy use in genomically low-risk disease. Importantly, however, their applicability and clinical confidence are reduced in lymph node–positive (LN+) disease, particularly in patients with ≥4 positive nodes and premenopausal patients with N1 disease, where contemporary guidelines explicitly qualify when these tests should and should not be used to guide chemotherapy decisions.

A major barrier to precision oncology is that most transcriptome-based biomarkers are discovered as outcome-associated gene lists rather than as readouts of the stable cellular programs tumors inherit from their cell-of-origin. Tissue identity and lineage programs shape epigenetic state, transcriptional wiring, and therapeutic vulnerabilities, and therefore provide a principled foundation for biomarker design. In contrast, widely used breast cancer assays were developed empirically from tumor expression and outcome, producing risk scores that are often difficult to interpret biologically and, in some settings, yield discordant risk assignments across assays. Our work is motivated by the premise that predicting the behavior of complex biological systems requires attention to initial conditions (7,8). In cancer, the relevant initial condition is the normal cell state from which the tumor arose. This framing is supported by evidence from systems biology and evolutionary/ecological dynamics showing that small differences in starting states can lead to divergent trajectories over time (9). A complementary principle is that unconstrained high-dimensional signatures can appear significant yet remain unstable across cohorts and resampling, motivating parsimonious models anchored in defined biology rather than purely data-driven correlation (10,11). Based on these two first principles, we set out to define breast cancer cell-of-origins with the fewest possible parameters.

Within this framework, we previously reported that breast cancer cell-of-origin determines metastatic potential and drug response of breast cancer in experimental models (12–14). We then found that a breast-specific triple-hormone receptor (THR) state, defined by the co-expression of estrogen (ER), androgen (AR), and vitamin D (VDR) receptors, delineates putative cells-of-origin with distinct epigenetic DNA methylation profiles (15,16). Finally, we reduced the THR cell-of-origin signature comprised of thousands of genes down to a 70-mRNA signature (THR-70) that proved to be prognostic across breast cancer subtypes (17). This signature was validated through outcome analyses spanning thousands of patients across publicly available cohorts and was further supported by single-cell evidence demonstrating enrichment in normal breast epithelium, consistent with a cell-of-origin program that is partially retained during transformation (17).

Here, we extend this platform by refining THR-70 into a minimal, clinically tractable signature that preserves prognostic information while enabling evaluation of treatment-response prediction in clinically relevant subgroups, including ER+/HER2− LN+ disease where current decision-making remains uncertain.

## RESULTS

### Interactome-guided refinement of the THR-70 cell-of-origin program identifies a six-gene core signature

To develop a clinically tractable cell-of-origin biomarker while preserving the biological structure of the previously reported THR-70 program (17), we applied an interactome-guided refinement strategy rather than purely statistical feature reduction. We reasoned that a minimal signature should retain coordinated expression, network connectivity, and lineage specificity, rather than maximizing prognostic signal through proliferation-driven gene selection.

Hierarchical clustering of THR-70 expression across ER+ breast cancers revealed five reproducible gene clusters (GCs) with distinct expression patterns (**Figure 1a**). These clusters segregated tumors into biologically interpretable expression states rather than reflecting a single continuous axis. Among them, one cluster (GC-4) exhibited the widest dynamic range across ER+ tumors, spanning low- and high-expression states across multiple intrinsic subgroups. To assess whether this cluster represented a coherent biological module, we evaluated protein–protein interaction structure using *STRING* (18). Genes within GC-4 formed a densely connected, high-confidence interaction network with significantly greater connectivity than expected by chance (**Figure 1b**), supporting the interpretation that these genes participate in a shared regulatory program. In contrast, other THR-70 clusters showed weaker or more fragmented interaction structure.

**Figure 1.**
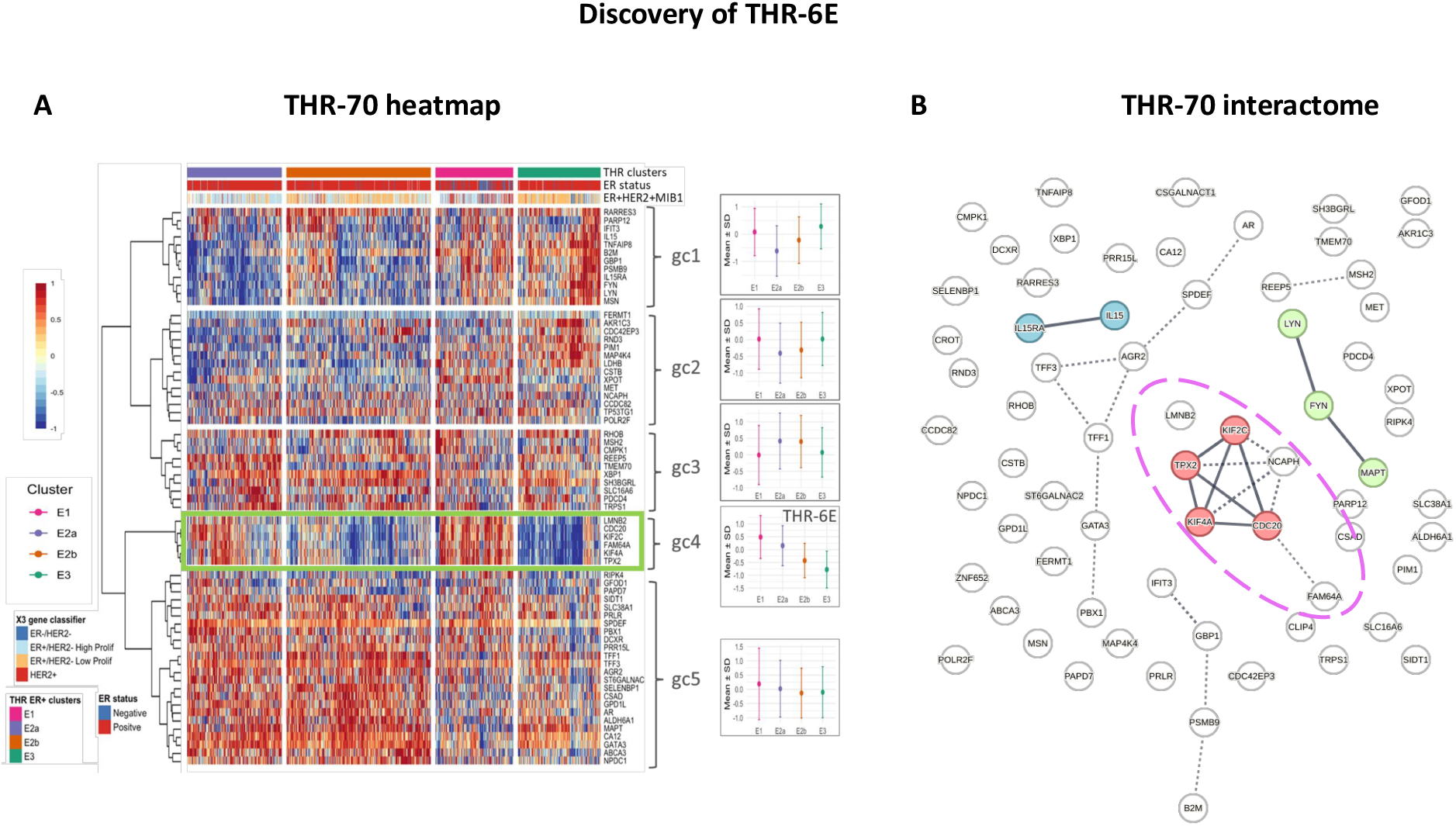
Interactome-guided refinement of THR-70 identifies the THR-6E module. **(a)** Heatmap of THR-70 expression across ER-positive breast cancers in the METABRIC cohort. Hierarchical clustering of ER-positive tumors defined four ER-positive tumor clusters (E1, E2a, E2b, E3). Genes in the THR-70 signature were partitioned into five gene clusters (gc1–gc5) based on their co-expression patterns across these tumor clusters. The THR-6E module corresponds to gene cluster 4 (gc4), which exhibits the greatest expression dynamic range across ER-positive tumors. The right panels summarize the distribution of expression for each gene cluster (gc1–gc5) across ER-positive tumors, shown as mean (point) and standard deviation (line). **(b)** Protein–protein interaction network of THR-70 genes generated using STRING (v12.0). Edges denote high-confidence interactions (combined score ≥0.70). Node groupings reflect cluster structure used for network visualization.

Across independent datasets, GC-4 genes were consistently over-expressed in tumors classified as high risk by recurrence outcomes (**Figure 2a and S1**), indicating that this module concentrates prognostically relevant information contained within the larger THR-70 program. We refer to this six-transcript signature as THR-6E, composed of Kinesin Family Member 4A and 2C (*KIF4A* and *KIF2C*) (19,20), Cell Division Cycle Protein 20 (*CDC20*) (21), Family With Sequence Similarity 64-Member A (*FAM64A*/*PIMREG*) (22), Microtubule Nucleation Factor (*TPX2*/*P100*) (23), and Lamin B2 (*LMNB2*) (24).

**Figure 2.**
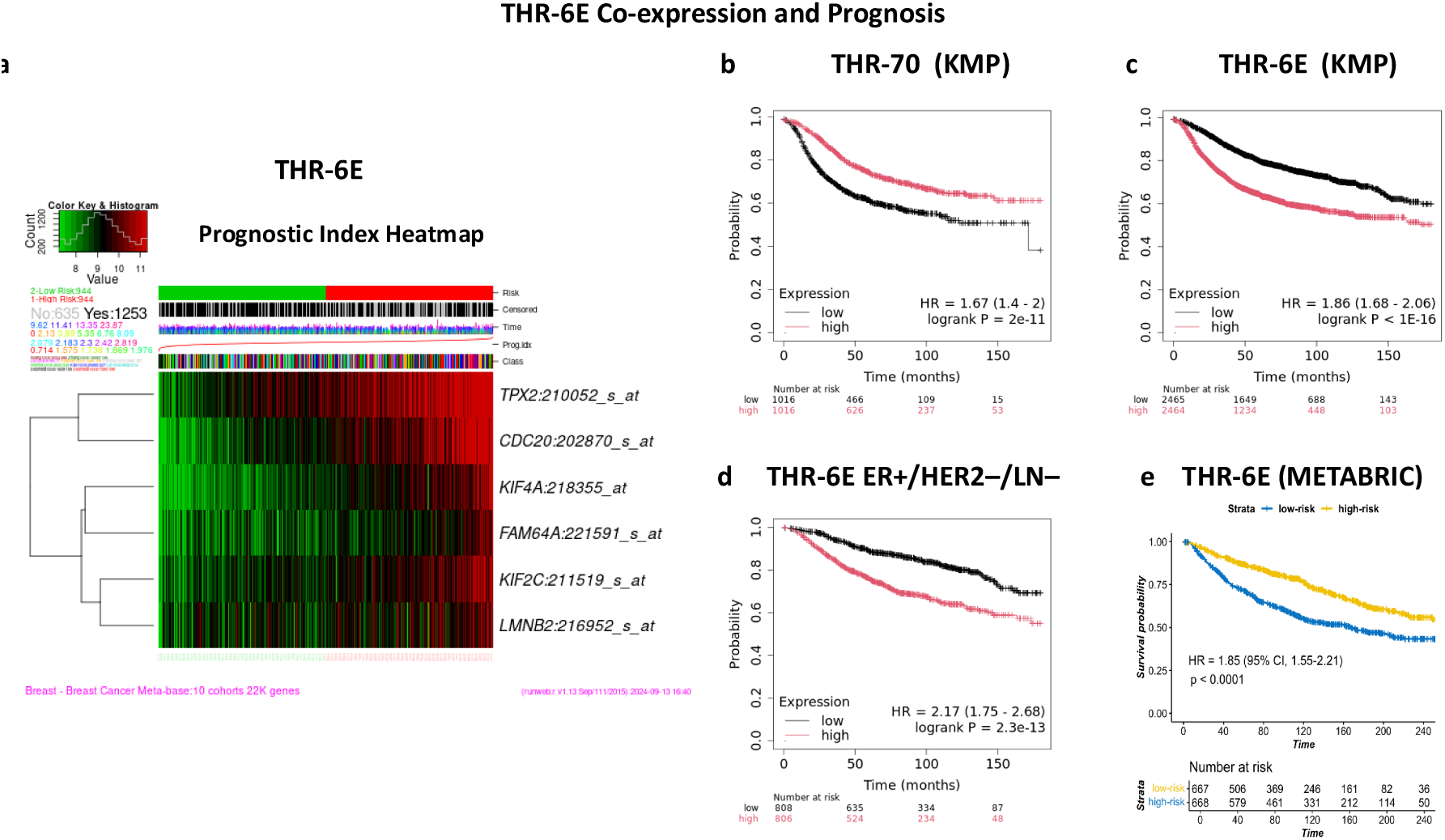
THR-6E preserves THR-70 prognostic performance. (**a**) SurvExpress risk-group heatmaps showing expression of THR-6E in the METABASE-10 breast cancer meta-cohort. Patients are stratified into low-risk (green) and high-risk (red) groups based on recurrence-free survival (RFS). (**b-e**) Kaplan-Meier plots for relapse-free survival (RFS) in the KMP cohort, comparing patients with high versus low mean expression using a median split. (**b**) THR-70 (RHR=1.67, p=2×10⁻¹¹). (**c**) THR-6E (RHR=1.84, p<1×10⁻¹⁶). (**d**) THR-6E in ER-positive (IHC), HER2-negative (array), lymph node-negative (ER+/HER2–/LN–) tumors in the KMP cohort (relative hazard ratio [RHR]=2.16, p=4.8×10⁻¹¹). (**e**) THR-6E in ER+/HER2–/LN– tumors in METABRIC. P-values are from two-sided log-rank tests; RHRs are from Cox proportional hazards models.

This refinement reduced the original THR-70 to a more compact six-gene signature while preserving a tightly coordinated, biologically interpretable core linked to luminal epithelial cell state. Notably, the genes comprising THR-6E are not selected on the basis of individual prognostic strength alone, but rather represent a compact, network-anchored module that captures the dominant cell-of-origin signal embedded within the parental THR program.

### THR-6E preserves the prognostic performance of THR-70 and stratifies risk in clinically relevant ER+/HER2− disease

We next tested whether compressing the THR program from 70 genes to six genes altered its ability to stratify clinical outcomes. Using the Kaplan–Meier Plotter (KMP) breast cancer collection, we compared relapse-free survival (RFS) separation achieved by THR-6E against the parent THR-70 signature using the same scoring framework (mean expression across genes) and the same patient stratification rule (median split) (**Figure 2b–c**). Despite its smaller size, THR-6E preserved prognostic performance, yielding an RFS relative hazard ratio (RHR) of 1.84 (p<1×10⁻¹⁶), outperforming THR-70 (RHR=1.67, p=2×10⁻¹¹), which reflects concentration of the prognostically informative THR signal into a compact module.

Because existing multigene assays are most commonly used in ER+/HER2–/LN– disease, we next examined THR-6E performance in this subgroup within the same KMP resource. In this subset, THR-6E robustly stratified RFS with an RHR of 2.16 (p=4.8×10⁻¹¹) (**Figure 2d**), indicating strong prognostic discrimination in the population where genomic testing is routinely deployed to inform adjuvant management decisions. Consistent with a breast lineage–anchored program, THR-6E showed limited or inconsistent prognostic association outside ER+/HER2− breast cancer, including ER−/HER2− and HER2+ breast cancers and selected non-breast tumor types (**Figure S2**).

To further determine whether these results generalize across cohorts, we validated THR-6E in the independent METABRIC cohort (25). Consistent with KMP, Kaplan–Meier analysis demonstrated significantly shorter RFS in the THR-6E high-risk group, with an RHR of 1.85 (p=6.0×10⁻¹²) (**Figure 2e**). The concordance of effect size and direction across KMP and METABRIC supports robustness to cohort composition and platform differences and establishes THR-6E as a reproducible prognostic signature in ER-positive breast cancer.

### THR-6E reflects a breast-specific luminal epithelial cell-of-origin program

A central characteristic of a cell-of-origin signature is tissue and cell-type specificity that is also preserved in malignant transformation and mapping to the normal epithelial lineage state from which the tumor arises. We have previously demonstrated, through single-cell transcriptomics, that THR-70 is enriched in normal breast epithelium (17). We therefore asked whether THR-6E exhibits coordinated breast specificity across orthogonal reference resources, rather than representing a broadly expressed proliferation module.

We first examined expression patterns across normal human tissues using ProteomicsDB (26). While several THR-6E genes show detectable expression in multiple organs when considered individually, their co-expression as a coherent module was most evident in breast tissue, particularly in the setting of ER, AR, and VDR expression (**Table 1**; **Figure S3a**). Specifically, breast tissue was the only organ in which ER/AR/VDR and all six THR-6E components were jointly present, whereas other tissues showed partial receptor expression with incomplete representation of the THR-6E module.

**Table 1.** Tissue and cell-specific expression of THR-6E genes. (a) Expression of breast-specific hormone receptors (ER, AR, and VDR) and THR-6E genes of Kinesin Family Member 4A and 2C (KIF4A, and KIF2C), Cell Division Cycle Protein 20 (CDC20), Microtubule Nucleation Factor (TPX2/P100), Family With Sequence Similarity 64, Member A (FAM64A/PIMREG) and Lamin B2 (LMNB2) in different normal human tissues.

| A. Tissue | ER-α | AR | VDR | KIF2C | KIF4A | CDC20 | FAM64A | TPX2 | LMNB2 |
| --- | --- | --- | --- | --- | --- | --- | --- | --- | --- |
| Breast | + | + | + | + | + | + | + | + | + |
| Cervix | + | + | + | - | - | - | - | - | + |
| Endometrium | + | + | - | - | + | - | - | - | + |
| Oviduct | + | + | - | - | - | - | - | - | + |
| Vagina | + | - | - | - | - | - | - | - | + |
| Liver | + | + | - | + | + | - | - | + | + |
| Kidney | - | + | + | - | + | - | - | - | + |
| Prostate | - | + | + | - | - | - | - | + | + |
| Gastrointestinal | - | - | + | + | + | - | - | + | + |
| Lung | - | - | + | + | + | + | - | + | + |
| Thyroid | - | - | + | - | - | - | - | - | + |
| Bladder | - | - | - | - | - | + | - | + | + |
| B. Breast Cells | ER-α | AR | VDR | KIF2C | KIF4A | CDC20 | FAM64A | TPX2 | LMNB2 |
| c-17 | + | + | + | - | - | - | - | - | + |
| c-21 | + | + | + | - | - | - | - | - | + |
| c-1 | + | + | + | - | + | + | - | - | - |
| c-16 | + | - | + | - | - | + | - | - | - |
| c-11 | + | - | + | - | - | + | + | - | - |
| c-20 | + | - | + | - | - | + | + | - | - |
| c-4 | + | - | + | - | - | + | + | - | + |
| c-9 | + | - | + | + | + | + | + | + | + |

To resolve THR-6E at cellular resolution within the normal breast, we analyzed the Human Protein Atlas single-cell transcriptomic dataset (27,28). THR-6E expression localized to glandular breast epithelial populations and exhibited a graded distribution across epithelial subsets (**Figure 3a** and **S3b**). A distinct epithelial cluster (C9) expressed all six THR-6E transcripts, whereas other glandular epithelial clusters expressed only subsets of the module (**Table 1**), indicating that THR-6E captures an epithelial cell-state spectrum rather than a binary marker of epithelial identity.

**Figure 3.**
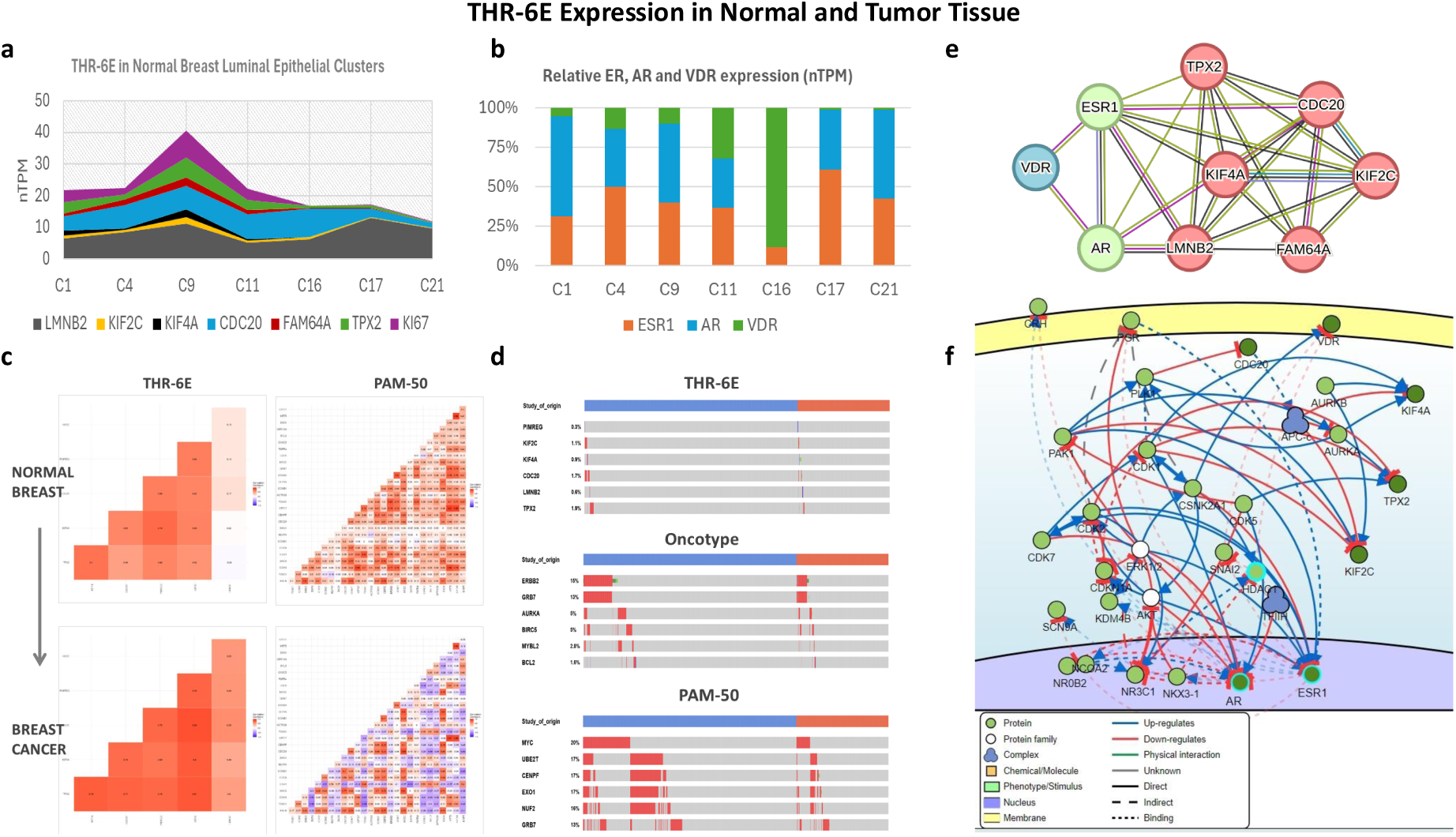
Analysis of THR-6E expression, mutation rates, and interactome in breast cancer. **(a)** THR-6E and MKI67 (Ki67) expression across normal breast glandular epithelial cell subsets in the Human Protein Atlas (HPA) single-cell RNA-seq dataset (v23). Values are reported as normalized transcripts per million (nTPM) for the indicated epithelial clusters (C1, C4, C9, C11, C16, C17, C21). **(b)** Fraction of cells expressing ESR1 (ER), AR, and VDR across the same glandular breast epithelial clusters in HPA (v23). Percent expression denotes the proportion of cells within each cluster with detectable transcript. **(c)** Gene–gene correlation matrices for THR-6E and PAM50 gene sets in normal breast tissue (top) and breast tumors (bottom) from TNMplot. Colors represent Spearman correlation coefficients (−1 to 1). **(d)** Genomic alteration frequencies for THR-6E, Oncotype DX, and PAM-50 gene sets in breast cancer using combined TCGA-BRCA and METABRIC cohorts (total n=3,593). Alterations include mutations and copy-number changes as reported by cBioPortal. THR-6E genes show low alteration frequency (mean 1.1%), whereas Oncotype DX and PAM-50 include multiple genes with recurrent copy-number gains (15–20% in queried genes). **(e)** Protein–protein interaction network of THR-6E with ESR1, AR, and VDR generated using STRING (v12.0) with k-means clustering. Orange arrows indicate literature-supported regulatory relationships overlaid on the STRING network. **(f)** CancerGeneNet (SIGNOR) network linking THR-6E genes to cancer-associated phenotypes. Query proteins are shown in yellow, first neighbors in green; protein families are shown as white circles and protein complexes as blue clover symbols. Solid edges denote direct interactions and dashed edges denote indirect interactions; blue arrows indicate up-regulation and red T-bars indicate down-regulation.

Notably, THR-6E-high epithelial clusters were enriched for proliferative features, with higher Ki67 expression observed in clusters C9, C1, C4, and C11 (**Figure 3a** and **S3b**). In contrast, clusters with strong ER and AR expression (C17 and C21) or high VDR expression (C16) showed comparatively low Ki67 levels (**Figure 3b**). This structure is consistent with a luminal epithelial differentiation axis in which hormone receptor–defined states and proliferative states are related but not interchangeable. Indeed, THR-6E is most strongly expressed in specific proliferative epithelial subsets, but it remains embedded within the broader THR-defined luminal lineage context rather than simply reflecting proliferation across all cell types.

Finally, we confirmed that THR-6E genes are expressed at the protein level in normal breast luminal epithelium (**Figure S4a**) and in breast cancer specimens (**Figure S4b**) using Human Protein Atlas immunohistochemistry (IHC). Together, the convergence of proteomic tissue profiling, single-cell transcriptomic localization, and tissue-based protein validation supports the conclusion that THR-6E is a breast-enriched luminal epithelial program consistent with a cell-of-origin signature, rather than a nonspecific proliferation surrogate.

### THR-6E expression patterns are preserved in breast cancer and enriched in metastatic disease

We then evaluated whether the coordinated THR-6E program observed in normal breast epithelium is maintained in breast tumors by comparing gene–gene correlation structure in normal and malignant tissues using TNMplot (29). THR-6E displayed a strikingly similar correlation matrix in normal breast tissue and breast cancer, indicating that its constituent genes remain tightly co-regulated after transformation (**Figures 3c** and **S5a**). In contrast, the correlation structure of Oncotype DX and PAM-50 gene sets differed substantially between normal and tumor tissues (**Figures 3c** and **S5a**). This divergence is expected for empirically derived tumor signatures, whose components reflect tumor-acquired biology, treatment-related selection, and stromal admixture rather than lineage-inherited programs. The preservation of THR-6E co-expression structure therefore supports its interpretation as a retained luminal cell-state module.

We next examined whether THR-6E is genetically stable in breast cancer, as would be predicted for a lineage-associated program not driven by recurrent genomic alteration. Using cBioPortal analyses of TCGA and METABRIC (3,593 total cases), THR-6E genes showed a low alteration frequency, averaging 1.1% across the six genes (**Figures 3d** and **S6**). By contrast, both Oncotype DX and PAM-50 gene sets included multiple genes with frequent copy number gains or other alterations in 15–20% of tumors (**Figures 3d** and **S6**). This contrast is important for clinical translation, since genomic instability within a signature gene set can distort expression-based scores over time, whereas the low alteration burden of THR-6E is consistent with a stable cell-of-origin readout less vulnerable to tumor evolutionary drift.

Finally, we tested whether THR-6E expression is breast-enriched at the pan-cancer level and whether it changes with metastatic progression. Pan-cancer analyses showed that THR-6E expression is highest in breast cancer and tracks the co-expression of ER, AR, and VDR (**Figure S7a**), consistent with its THR lineage foundation. Within breast tissue specifically, THR-6E expression was significantly increased in metastatic lesions compared with primary tumors (p=1.7×10⁻⁵⁴; **Figure S7b–d**). The directionality of this shift suggests that the THR-6E program is not diluted during progression; rather, it becomes further accentuated in metastasis.

### THR-6E forms a hormone-linked regulatory network connected to proliferation and epithelial biology

Having established that THR-6E represents a coherent, breast-enriched epithelial program that is preserved in tumors, we next asked whether its components connect mechanistically to the hormone receptor axis that motivated the original THR framework. First, we examined protein–protein interaction structure by jointly querying THR-6E components with ESR1 (ER), AR, and VDR. This analysis revealed a significantly enriched interaction network, with more observed interactions than expected under the null model (PPI enrichment p=1.86×10⁻⁷) (**Figures 3e** and **S8a**). The resulting network places THR-6E proteins in close proximity to hormone receptor signaling, supporting the interpretation that the six-gene module captures an epithelial program that is functionally coupled to the THR axis.

To complement network topology with published mechanistic relationships, we curated experimentally supported links between THR-6E genes and hormone receptor signaling. Prior studies have shown that ER and AR can transcriptionally upregulate KIF4A (19,30), and that KIF4A can bind AR and prevents its degradation (19), providing a plausible feedback linkage between cytoskeletal motor programs and androgen signaling. AR upregulation of FAM64A/PIMREG further supports direct coupling between androgen signaling and THR-6E components (22). In addition, TPX2 has been reported as an AR co-activator (23), providing a mechanistic bridge between microtubule/spindle-associated machinery and hormone-driven transcriptional control. Finally, Lamin-associated regulation of ER expression (24) links nuclear architecture to hormone receptor signaling, consistent with a differentiation-linked epithelial state program rather than an isolated proliferation module (**Figures 3e** and **S8b**).

We then asked what cancer-relevant phenotypes are most plausibly downstream of THR-6E biology. Using *CancerGeneNet* (31), we mapped causal and regulatory relationships linking THR-6E genes to hallmark phenotypes. This analysis positioned THR-6E within a breast-relevant network enriched for hormonal regulation and epithelial biology, with connections to spindle assembly and microtubule organization, intracellular transport, Golgi–ER membrane trafficking, survival, and proliferation (**Figures 3f** and **S8b**).

Taken together, these network analyses support a mechanistic interpretation of THR-6E as a hormone-linked epithelial program that interfaces with core proliferation machinery while retaining features of luminal cell-state biology. This network structure provides biological plausibility for why THR-6E, derived from a THR cell-of-origin framework, concentrates prognostic and treatment-response information in ER-positive breast cancer.

### THR-6E predicts systemic therapy outcomes across grade, subtype, and nodal status

We next evaluated the clinical performance of THR-6E in ER+/HER2– breast cancer treated with systemic therapy to determine whether the signature remains informative across clinically relevant strata defined by grade, intrinsic subtype, and lymph node status. Using the KMP resource, we first examined ER+/HER2− tumors receiving any systemic therapy. Across 3,201 treated patients, high THR-6E expression identified a group with significantly shorter RFS, with a hazard ratio of 2.05 (p=1.0×10⁻¹⁶) using the median split and 2.12 (p=1.0×10⁻¹⁶) using the optimized cutoff (**Figure 4a**; **Table 2**).

**Figure 4.**
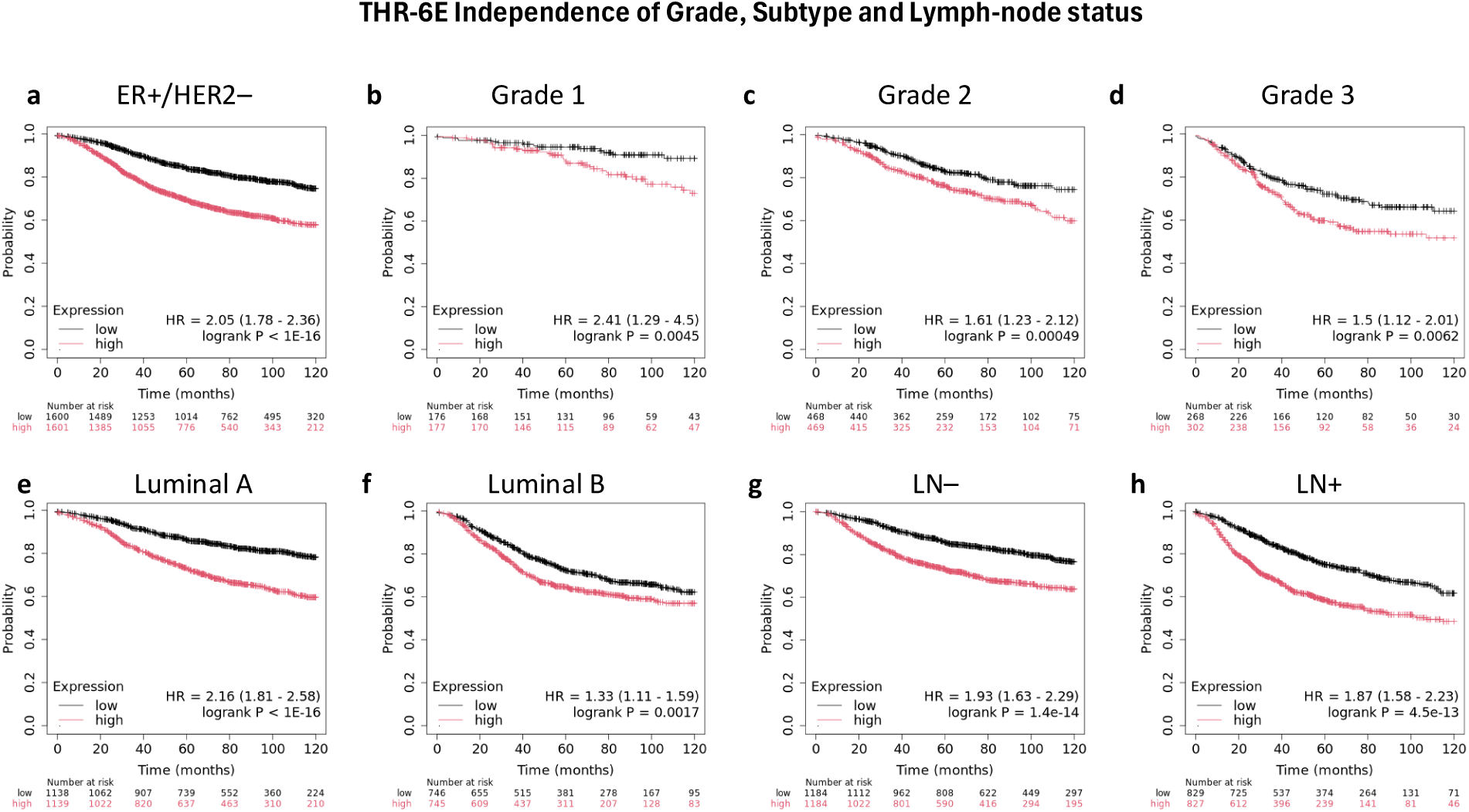
THR-6E stratifies relapse-free survival among systemically treated ER+/HER2− breast cancer patients across clinical subgroups. **(a)** Kaplan–Meier relapse-free survival (RFS) curves in ER+/HER2– tumors stratified by THR-6E expression using median cutoff. **(b-d)** RFS curves in systemically treated ER+/HER2− tumors stratified by THR-6E in grade 1 **(b)**, grade 2 **(c)**, and grade 3 **(d)** tumors. **(e-f)** RFS curves in Luminal A **(e)** and Luminal B **(f)** tumors that were treated with systemic therapy, stratified by THR-6E expression. **(g-h)** Kaplan–Meier RFS curves in lymph node-negative **(g)** and lymph node-positive **(h)** breast cancer patients treated with any systemic therapy, stratified by THR-6E expression. For all panels, patients were dichotomized into THR-6E high and low groups using the median cutoff of mean THR-6E expression. Hazard ratios (HR) with 95% confidence intervals (CI) and two-sided log-rank p-values are shown on each plot; numbers at risk are displayed below curves.

**Table 2.** Prognostic and Predictive Comparison of Signatures. The relative hazard ratio (RHR) and 95% confidence interval (CI) for relapse-free survival (RFS) are analyzed in ER+/HER2– breast cancers using the Kaplan-Meier Plotter (KMP), as well as in estrogen-positive, HER2-negative (ERpHER2n) breast cancers from the METABRIC datasets. The response to endocrine therapy in ERpHER2nLNn and the response to chemotherapy in ERpHER2nLNp breast cancers are assessed using the area under the curve (AUC). Results that are not statistically significant (p > 0.5) are highlighted as NS. (*) Oncotype relative hazard ratio (RHR) is inverted for comparison with THR-6E.

| ER+/HER2- breast cancer |  | Median |  |  | Optimum cut-off |  |
| --- | --- | --- | --- | --- | --- | --- |
| Subtype | Treatment | N | HR | P-value | HR | P-value |
| ER+/HER2– | Systemic therapy | 3,201 | 2.05 | 1.0E-16 | 2.12 | 1.0E-16 |
|  | Hormone therapy | 789 | 1.99 | 7.0E-06 | 2.31 | 8.0E-09 |
|  | Chemotherapy | 281 | 1.76 | 1.4E-02 | 3.53 | 3.1E-04 |
| Luminal A | Systemic therapy | 2,277 | 2.16 | 1.0E-16 | 2.20 | 1.0E-16 |
| Luminal B |  | 1,491 | 1.33 | 1.7E-03 | 1.48 | 2.4E-05 |
| Grade1 | Systemic therapy | 353 | 2.41 | 4.5E-03 | 2.84 | 2.0E-04 |
| Grade2 |  | 936 | 1.61 | 4.9E-04 | 2.03 | 2.5E-07 |
| Grade3 |  | 570 | 1.31 | 6.8E-02 | 1.50 | 6.2E-03 |
| LN– | Systemic therapy | 2,368 | 1.93 | 1.4E-14 | 1.96 | 4.0E-15 |
|  | Hormone therapy | 540 | 1.76 | 5.7E-04 | 2.62 | 7.8E-07 |
| LN+ | Systemic therapy | 1,656 | 1.87 | 4.5E-13 | 2.45 | 1.6E-15 |
|  | Chemotherapy | 799 | 1.62 | 2.0E-04 | 2.12 | 1.2E-07 |

We then asked whether the association persisted within standard clinicopathologic strata that often dominate adjuvant treatment decisions. When stratified by histologic grade among systemically treated ER+/HER2− tumors, THR-6E remained prognostic across grades. Using the optimized cutoff, hazard ratios were 2.84 for grade 1 (p=2.0×10⁻⁴), 2.03 for grade 2 (p=2.5×10⁻⁷), and 1.50 for grade 3 (p=6.2×10⁻³) (**Figure 4b–f**; **Table 2**). The persistence of effect across grades argues against THR-6E functioning simply as a surrogate for grade-associated proliferation and suggests it captures additional biology relevant to outcomes under treatment. In luminal subtypes, THR-6E stratified outcomes in Luminal A tumors, with hazard ratios of 2.16 (p=1.0×10⁻¹⁶) using the median split and 2.20 (p=1.0×10⁻¹⁶) using the optimized cutoff. In Luminal B tumors, THR-6E also remained significant, with hazard ratios of 1.33 (p=1.7×10⁻³) and 1.48 (p=2.4×10⁻⁵), respectively (**Figure 4i–j**; **Table 2**).

Finally, we evaluated performance across lymph node status, given the known limitations of existing genomic assays in node-positive disease. In patients with ER+/HER2−/LN– tumors receiving systemic therapy (n=2,368), THR-6E stratified RFS with a hazard ratio of 1.93 (p=1.4×10⁻¹⁴) using the median split and 1.96 (p=4.0×10⁻¹⁵) using the optimized cutoff (**Figure 4g**; **Table 2**). Notably, among LN+ treated patients (n=1,656), THR-6E remained strongly associated with outcome, with hazard ratios of 1.87 (p=4.5×10⁻¹³) and 2.45 (p=1.6×10⁻¹⁵) under the median and optimized cutoffs, respectively (**Figure 4h**; **Table 2**). These findings are particularly important given that several established signatures are not recommended by the ASCO or NCCN for breast cancers involving more >3 positive LNs (32).

### THR-6E predicts endocrine and chemotherapy response, including in high-grade and node-positive disease

We next asked whether THR-6E provides predictive information for treatment response, rather than only stratifying prognosis. Because ER+/HER2− breast cancer is treated with both endocrine therapy and chemotherapy depending on risk and nodal status, we evaluated THR-6E in treatment-response–annotated cohorts using RFS, response in endocrine-treated disease, and pathologic complete response (pCR) in chemotherapy-treated disease.

First, we assessed whether THR-6E discriminates responders from non-responders to endocrine therapy among ER+/HER2–/LN– patients. THR-6E achieved an AUC of 0.63 (p=1.2×10⁻⁷) (**Figure 5a**). Consistent with the directionality observed in survival analyses, higher THR-6E expression was enriched in endocrine non-responders, with a 1.2-fold increase compared with responders (p=1.3×10⁻⁶) (fig. 5a). These results support an association between elevated THR-6E expression and reduced endocrine sensitivity, rather than simply reflecting baseline prognosis.

**Figure 5.**
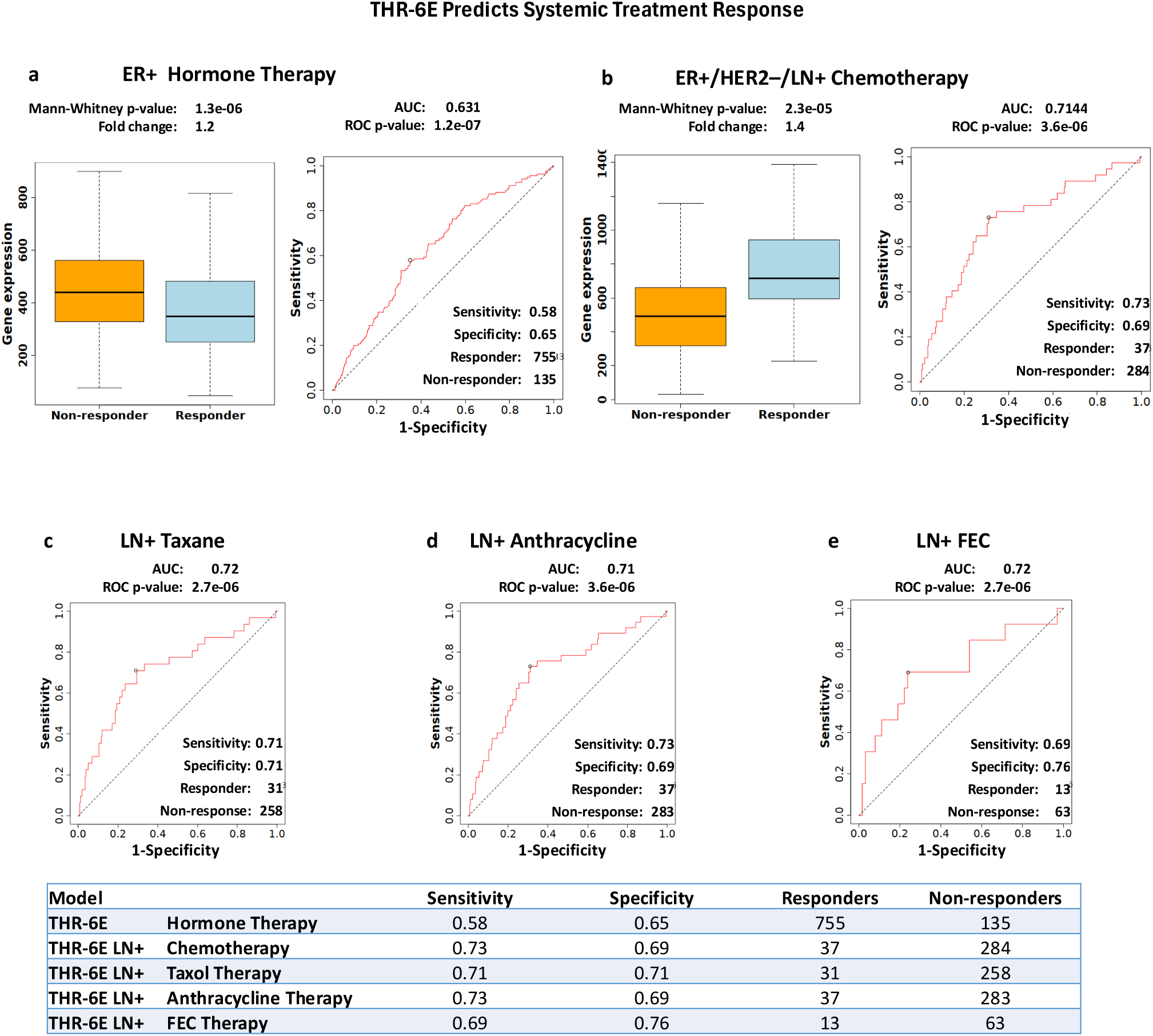
THR-6E predicts endocrine-therapy and chemotherapy response in ER-positive breast cancer. For each panel, the top boxplots compare THR-6E signature expression between responders and non-responders, and the bottom panels show receiver operating characteristic (ROC) curves summarizing classification performance. Predictive accuracy is reported as area under the ROC curve (AUC), sensitivity, and specificity. (**a**) Endocrine therapy response in ER-positive, lymph node–negative (ER+/LN–) breast cancer. (**b–e**) Chemotherapy response in ER+/HER2–/LN+ breast cancer: all chemotherapy (**b**), taxane-based regimens (**c**), anthracycline-based regimens (**d**), and FEC (fluorouracil, epirubicin, cyclophosphamide) regimens (**e**).

We then evaluated chemotherapy response in ER+/HER2–/LN+ patients using pCR as the endpoint. In this setting, THR-6E demonstrated balanced predictive performance with approximately 70% sensitivity and specificity, achieving an AUC of 0.71 (p=3.6×10⁻⁶) (**Figure 5b**). Importantly, performance was consistent across major chemotherapy regimen classes, supporting that the signal is not restricted to a single drug backbone. Specifically, AUC was 0.72 for taxane-based therapy (p=2.7×10⁻⁶), 0.71 for anthracycline-based therapy (p=3.6×10⁻⁶), and 0.72 for FEC (p=2.7×10⁻⁶) (**Figure 5c–e**).

Because grade III tumors are often treated aggressively and can exhibit heterogeneous response behavior despite uniformly high-risk histology, we examined whether THR-6E retains predictive value within high-grade disease. THR-6E remained predictive of chemotherapy response among grade III tumors, including those with LN+ status (**Figure S9**), further supporting its potential role as a biomarker that refines treatment expectation within groups where current clinical variables provide limited resolution.

### Independent validation in the I-SPY2 trial demonstrates treatment-specific predictive performance

To evaluate THR-6E in a trial-grade setting with standardized therapy delivery and centrally adjudicated response endpoints, we performed independent testing in the I-SPY2 neoadjuvant clinical trial (33,34), focusing on patients with ER+/HER2– tumors (n=379; 64 pCR). Across all treatment arms combined, the THR-6E model achieved an AUC of 0.74, with sensitivity of 0.68 and specificity of 0.71 (**Figure 6a**).

**Figure 6.**
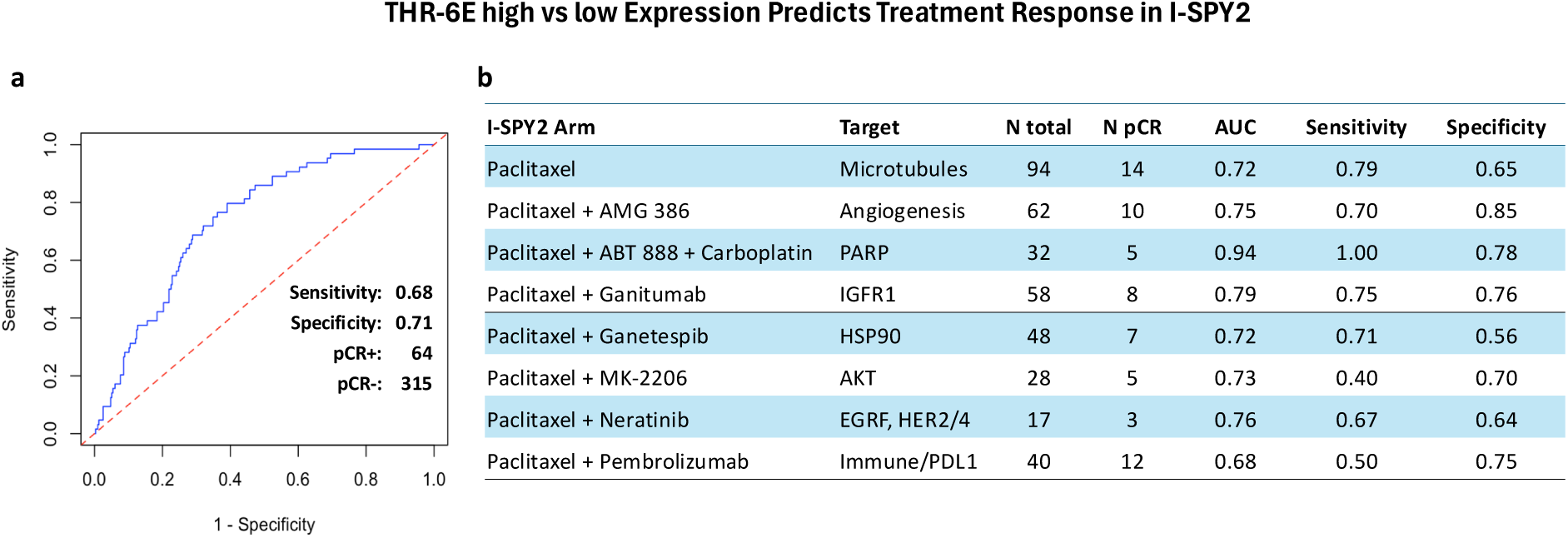
Independent validation in I-SPY2 demonstrates treatment-arm–specific prediction of pathologic complete response. **(a)** Receiver operating characteristic (ROC) curve for predicting pathologic complete response (pCR) in the ER+ /HER2– I-SPY2 cohort. A linear SVM classifier was trained on external pCR-annotated datasets and evaluated in I-SPY2 without refitting. Overall performance across all I-SPY2 treatment arms combined is shown (AUC=0.741; sensitivity=0.68; specificity=0.71). **(b)** Arm-level performance summary in I-SPY2, reporting therapeutic target, number of patients per arm (N), number achieving pCR, and corresponding AUC, sensitivity, and specificity values for each treatment arm.

When analyzed by treatment arm, predictive performance varied substantially, consistent with drug-mechanism dependence and heterogeneous response biology across regimens (**Figure 6b**). The strongest discrimination was observed in the paclitaxel + ABT-888 (veliparib) + carboplatin arm, where the model achieved an AUC of 0.94 with sensitivity of 1.00 and specificity of 0.78. Several targeted therapy arms also demonstrated robust performance, including paclitaxel + ganitumab (AUC 0.79), paclitaxel + neratinib (AUC 0.76), and paclitaxel + AMG 386 (AUC 0.75). In contrast, the immune-based paclitaxel + pembrolizumab arm showed more modest discrimination (AUC 0.68), consistent with greater biological heterogeneity in response determinants in immunotherapy-containing regimens within ER+/HER2− disease.

## DISCUSSION

While both pre-existing (nature) and acquired (nurture) traits influence biology, cancer research has often prioritized acquired mutations over tissue-specific characteristics. However, a recent pan-cancer study involving 32 cancer types and 10,884 patients revealed that oncogenes and mutations are not necessary always prognostic (35), and that, on average, 46% of top-scoring prognostic features were related to gene expression, 20% to DNA methylation, only 1% to mutations (35). In this context, our findings support a cell-of-origin framework in which stable, lineage-inherited epithelial programs establish a baseline trajectory that shapes both recurrence risk and therapeutic sensitivity. Supporting this framework, Hoadley et al. underscored the significance of tissue origin in understanding cancer biology across various tumor types (36), and Polak et al. found that cell-of-origin chromatin organization also shapes the mutational landscape of cancer (37).

We have previously demonstrated that different normal breast cell origins are associated with distinct methylation and gene expression profiles (13,15,38); however, translating these genome-wide cell-of-origin patterns into clinical applications has proven challenging. Here, we introduce THR-6E, a compact, network-anchored refinement of a broader triple hormone receptor cell-of-origin program (17). Several properties of THR-6E align with expectations for a cell-of-origin signature. First, THR-6E localizes to glandular breast epithelium in single-cell reference data and shows tissue-restricted co-expression at the protein level, consistent with a luminal epithelial lineage program rather than a ubiquitous stress or cell-cycle readout. Second, the internal correlation structure of THR-6E is preserved between normal breast tissue and breast tumors, supporting inheritance and partial retention of a normal epithelial state during transformation. Third, THR-6E genes are rarely altered genomically in breast cancer, in contrast to empirically derived multigene panels that include recurrently amplified or otherwise altered driver genes. This distinction is important since signatures dominated by tumor-acquired genomic events can drift as tumors evolve under therapy and selection, whereas a lineage-anchored program is expected to be more stable across time and across cohorts.

Clinically, THR-6E preserved the prognostic performance of the parent THR-70 program and remained robust across independent datasets. Importantly, it stratified relapse-free survival within ER+/HER2− disease across grades and luminal subtypes and retained strong performance in LN+ disease, where data are insufficient to guide chemotherapy decisions particularly in patients with ≥4 positive nodes (32) and premenopausal N1 patients (39). These observations support the premise that a biology-anchored signature can complement existing tools by extending consistent risk stratification into clinically ambiguous settings that are frequently excluded from assay-driven de-escalation decisions.

Beyond prognosis, THR-6E carried predictive signal for systemic therapy response. In endocrine-treated ER+/HER2−, node-negative disease, higher THR-6E expression was enriched in non-responders and yielded statistically robust discrimination. In chemotherapy-treated ER+/HER2−, node-positive disease, THR-6E predicted pCR across multiple regimens in the I-SPY2 trial, which demonstrates generalization in a standardized, prospectively conducted setting. Notably, predictive performance varied across arms, with particularly strong discrimination in patients treated with Paclitaxel in combination with Ganetespib, Ganitumab, AMG386, and ABT 888 plus Carboplatin. However, performance was more modest in the pembrolizumab arm, which is expected given that THR-6E appears to capture an epithelial cell-state baseline that may interact more directly with cytotoxic and DNA-damage-linked vulnerabilities than with immune-mediated response determinants, which are shaped by additional microenvironmental and host factors. These findings support the use of THR-6E as a biomarker for treatment deescalation in ER+/HER2– disease, which aligns with the growing need for biomarkers guiding targeted therapies.

A recurring critique of expression-based biomarkers is that many signatures, including random gene sets, can correlate with outcome because proliferation is broadly coupled to prognosis in breast cancer (40). Our findings suggest that THR-6E captures a luminal epithelial program that exists in normal breast tissue and is conserved in breast cancer, independent of proliferation. First, THR-6E was derived from a cell-of-origin framework rather than from outcome-driven selection and forms a high-confidence interactome module linked to hormone receptor biology. Furthermore, it is localized to breast epithelial cell populations rather than broadly expressed across tissues. At the same time, the data also show that THR-6E intersects with proliferative epithelial states in normal breast epithelium, which is expected for a luminal epithelial program that includes regulated cell-cycle machinery.

Several translational implications follow. First, THR-6E offers a plausible route to expand molecular guidance for ER+/HER2−/LN+ patients, including those with higher nodal burden where assay guidance is limited (32). Second, the stability of the signature and its anchoring in cell-of-origin biology support integration with dynamic biomarkers such as circulating tumor DNA to model baseline risk together with evolving minimal residual disease. Third, because THR-6E is compact and composed of genes measurable across platforms, it is inherently more amenable to standardized implementation than large proprietary panels, provided that scoring rules and cutpoints are prespecified and analytically validated. Finally, the network connectivity of THR-6E to ER/AR/VDR biology raises testable therapeutic hypotheses, including whether THR-6E–defined states mark tumors more likely to benefit from pathway-targeted modulation in conjunction with standard endocrine or cytotoxic therapy.

This study has some inherent limitations. First, the analyses are retrospective and utilize publicly available data from several cohorts. While this enables breadth and independent replication, treatment annotations, dosing, and endpoint definitions are not perfectly harmonized across cohorts. The ROC-based analyses rely on available responder/non-responder labeling and therefore should be interpreted as evidence of predictive signal rather than definitive clinical decision thresholds. The I-SPY2 results provide strong external validation but remain limited by sample size within individual treatment arms, which can widen uncertainty around arm-specific estimates even when point estimates are compelling. In addition, while THR-6E is biologically grounded, this study does not establish causal mechanisms linking the signature to drug sensitivity; however, it establishes reproducible association patterns across cohorts, modalities, and a trial setting.

Importantly, hallmark enrichment analysis did not identify significant enrichment of canonical cancer hallmark programs within THR-6E, in contrast to Oncotype DX (**Figure S10**); therefore, combining THR-6E with signatures representing acquired hallmark modules (e.g., immune, metabolism, invasion, angiogenesis) could further improve predictive performance (41,42). Additionally, integrating THR-6E with computational tools like PREDICT or Adjuvant, which consider clinical variables like tumor grade and size, may significantly boost its overall effectiveness. Such strategies could potentially optimize the clinical utility of the THR-6E signature and lead to more targeted therapeutic approaches.

Taken together, our results support a model in which lineage-inherited epithelial state is a central determinant of both recurrence risk and therapeutic response in ER+/HER2− breast cancer. THR-6E distills breast cell-of-origin anchored in the co-expression of ER/AR/VDR into a six-gene signature that is specific to normal and malignant breast epithelium, genomically stable, and predictive across clinically relevant strata, including LN+ disease. More broadly, these findings align with emerging evidence that tissue and cell-of-origin remain clinically consequential even as pan-cancer, tissue-agnostic frameworks expand (36,36,37). They also underscore that therapeutic response is constrained by the cellular programs cancers inherit and retain and not solely as a function of acquired mutations. Prospective diagnostic accuracy studies and head-to-head comparisons against existing assays in pre-specified clinical contexts will be essential to define where THR-6E adds value, how it should be integrated into clinical decision-making, and whether THR-6E-guided strategies improve patient outcomes.

## METHODS

### Samples and Cohorts

Prognostic analyses were performed in two primary breast cancer cohorts: the Molecular Taxonomy of Breast Cancer International Consortium (METABRIC; n=1,904) (25) and the Kaplan–Meier Plotter breast cancer compendium (KM Plotter; n=2,032 across 50 studies) (43). Signature derivation and clustering to identify the THR-6E module were performed using ER-positive tumors in METABRIC, as shown in **Figure 1**. Treatment-response analyses were conducted using *ROCplot*, which aggregates transcriptomic data with endocrine-therapy and chemotherapy response annotations across multiple breast cancer studies (44), and were restricted to the clinically defined subgroups reported in the Results. Independent validation of pCR prediction was performed using the I-SPY2 ER+/HER2– cohort (n=379; 64 pCR), with model training conducted on an external training cohort assembled from eight independent GEO datasets with pCR annotations: GSE191127, GSE192341, GSE25066, GSE41998, GSE16716, GSE20271, GSE32646, and GSE34138 (combined n=754; 78 pCR).

### Development of the THR-6E Gene Signature

THR-6E was derived by refining the previously established 70-gene triple hormone receptor cell-of-origin signature (THR-70), which stratifies breast cancer into five groups, including four ER-positive clusters (E1, E2a, E2b, E3) (17). THR-70 genes were partitioned into five gene clusters based on expression patterns across ER-positive tumors. To identify a minimal module capturing the dominant dynamic range across ER-positive disease, expression variability within each gene cluster was quantified across the four ER-positive tumor groups by comparing summary statistics (mean and standard deviation) within each cluster across groups. The cluster exhibiting the greatest variability across ER-positive tumor groups was selected as the refined module, yielding the six-gene THR-6E signature.

### Gene Expression Analysis

Heatmaps comparing THR-6E and Oncotype gene expression across risk groups were generated using *SurvExpress* (45) on the METABASE-10 breast cancer dataset, which comprises ten different studies, nine of which overlap with the KMP cohort except for GSE4922 (n = 249). Analyses were run using *SurvExpress* default settings except where specified: survival days were not censored; samples were stratified by author; heatmap by risk was selected with two risk groups; network analysis was disabled; imputation and quantization were disabled; and advanced/attribute plot options were enabled. Risk-group separation p-values were computed using log-rank test, and relative hazards were estimated using Cox proportional hazard models. THR-6E expression in normal tissue, primary tumors, and metastatic samples was evaluated using TNMplot (29), which integrates gene expression data derived from NCBI-GEO, TCGA, Therapeutically Applicable Research to Generate Effective Treatments (TARGET), and the Genotype-Tissue Expression (GTEx). The referenced TNMplot resource includes 14,918 normal samples, 409,442 tumor samples, and 848 metastatic samples.

### Mutational Profiling and Copy-Number Alterations

Genomic alterations affecting THR-6E and comparator signatures in both TCGA breast invasive carcinoma (RNA Seq V2; 1,082 samples/patients) and the METABRIC (2,051 samples/patients) cohorts were retrieved from CBioPortal and analyzed. Alterations were visualized using the OncoPrint view with OncoKB and hotspot annotation options enabled, and the display restricted to profiled samples for the queried genes/profiles.

### Proteome Analysis

Protein-level expression of THR-6E components and hormone receptors was evaluated using ProteomicsDB (26) to assess tissue-level co-expression patterns. The following UniProt identifiers were queried: ER/ESR1 (P03372), AR (P10275), VDR (P11473), KIF2C (Q99661), KIF4A (O95239), CDC20 (Q12834), TPX2 (Q9ULW), PIMREG/FAM64A, and LMNB2 (Q03252). Immunohistochemistry for THR-6E gene products in normal breast and breast cancer tissues was assessed using the Human Protein Atlas portal (27,28).

### Interactome and Phenotype-Network Analyses

Protein–protein interaction structure was evaluated using STRING (v12.0) (18). THR-6E genes and hormone receptors (ESR1, AR, VDR) were entered using the “Multiple proteins” query. Network clustering was performed using k-means clustering with three clusters. Phenotype-linked network relationships were evaluated using CancerGeneNet (SIGNOR) (31). THR-6E genes and hormone receptors (ESR1, AR, VDR) were entered in the “connect proteins” interface. Analyses were performed using: interaction type = all; score = 0.0; layout = relaxed; toolbox complexity level = level 2 (first neighbors). CancerGeneNet returns a graph of causal and regulatory paths linking input genes to cancer-associated phenotypes.

### Survival Analysis

In the KMP cohort (n = 2,032, 50 studies), survival analyses were performed using the KM Plotter breast cancer interface (43). Unless otherwise specified, analyses used relapse-free survival as the endpoint; patients were split by median expression for multi-gene signatures; follow up threshold was set to all; and censoring at threshold was enabled. For multi-gene signatures, the “use multiple genes” option was selected and probe selection used the user-selected probe set/JetSet where applicable. The “invert HR values below 1” option was enabled for consistent directionality. Cohort filters were applied as relevant to each analysis (e.g., ER status by IHC, HER2 status by array, LN status, grade, and PAM-50 subtypes). Redundant samples and biased arrays were excluded from analysis and proportional hazards assumptions and multivariate analysis options were enabled where indicated (including MKI67, ESR1, and ERBB2/HER2 as covariates). The Benjamini-Hochberg method was used to correct for multiple hypothesis testing.

In METABRIC, a logistic regression model was used to compute a continuous risk score for each patient using the expression levels of the signature genes. Patients were subsequently categorized into low- and high-risk using optimum thresholds from the ROC curve of the training set.

### Treatment Response Analyses

For gene expression datasets available from the ROCplot resource (44), treatment response prediction analyses were performed using differentially expressed genes to classify tumor samples into responders vs. non-responders. Response type (relapse-free survival response or pathological response), treatment type (endocrine therapy or chemotherapy), and clinical filters (grade, nodal status, ER status, HER2 status, molecular subtype) were set as specified for each panel. JetSet-only probe selection and no-outlier filtering were enabled; quartile display was disabled. Predictive accuracy was summarized using AUC with corresponding p-values (44).

To evaluate THR-6E as a predictor of pCR in ER+/HER2– breast cancer treated with neoadjuvant chemotherapy, an external training set was constructed from eight independent cohorts with pCR annotation (GSE191127, GSE192341, GSE25066, GSE41998, GSE16716, GSE20271, GSE32646, GSE34138). Expression matrices were normalized and log-scaled prior to integration, followed by quantile normalization and removal of duplicate samples. The final training cohort included 754 ER+/HER2– patients, of whom 78 achieved pCR. Independent validation was performed in the I-SPY2 ER+/HER2– cohort (n=379; pCR in 64 patients). A linear support vector machine (SVM) classifier was trained on the external training cohort and evaluated on I-SPY2 without refitting. Model performance was reported as ROC AUC, sensitivity, and specificity across all treatment arms and within individual arms. The decision threshold for converting model outputs into binary predictions was selected using the training cohort and applied unchanged in I-SPY2 to preserve an independent test evaluation.

### Single-cell mRNA Expression Analysis

Normal breast single-cell transcriptomic analyses were performed on the GSE164898 dataset (46), available through the Human Protein Atlas (HPA) version 23.0. This dataset comprises gene expression data from 46,126 cells grouped into 557 individual cell type clusters corresponding to 15 different cell types. Briefly, this dataset was processed by HPA using an in-house Python pipeline (Scanpy v1.7.1). Cells were filtered for valid data, requiring at least 200 detected genes per cell and genes expressed in at least 10% of cells. Valid cells were clustered using Louvain clustering with default parameters, projecting features into PCA space with 50 components and generating a k-nearest neighbors (KNN) graph. Clusters were defined using 15 neighbors and a resolution of 1.0. Cluster-level expression was summarized as transcripts per million (pTPM) for visualization of THR-6E genes, ER/AR/VDR, and Ki67 across annotated glandular breast epithelial clusters.

### Software and Statistical Analysis

Unless otherwise specified, statistical analyses were conducted in R (v4.0.3). Kaplan–Meier curves and Cox proportional hazards models were generated using the survival and survminer packages. Hierarchical clustering used functions from the stats package, and logistic regression modeling used glmnet where regularized fitting was required. Two-sided statistical tests were used throughout, and significance was assessed at p<0.05 unless otherwise specified. Multiple hypothesis testing corrections were applied using Benjamini–Hochberg method. Survival curves were compared using the log-rank test.

## DATA AND CODE AVAILABILITY

The gene expression datasets used in this study are available through the Gene Expression Omnibus (GEO) using the accession numbers provided in the manuscript. All online resources used to conduct this study are publicly available and can be accessed at the following URLs: KMplot (https://kmplot.com/analysis/index.php?p=service&cancer=breast), ProteomicsDB (https://www.proteomicsdb.org/), the Human Protein Atlas (https://v23.proteinatlas.org/), cBioPortal (https://www.cbioportal.org/), TNMplot (https://tnmplot.com/analysis/), ROCplot (https://www.rocplot.com/site/treatment), STRING (https://string-db.org/), SIGNOR/CancerGeneNet (https://signor.uniroma2.it/CancerGeneNet/), and SurvivaX (https://victortrevino.bioinformatics.mx:8080/Biomatec/SurvivaX.jsp). The code required to reproduce the results presented in this study will be made publicly available on GitHub upon publication.

## ACKNOWLEDGMENTS

We thank Drs. Alexander Borowsky and Saurabh Mehta for their suggestions, which greatly improved the manuscript.

## AUTHOR CONTRIBUTIONS

T.A.I and M.O. conceived and supervised the project. I.V., P.V. conducted the analysis. T.A.I and M.O. wrote the manuscript draft. All authors read and approved the manuscript.

## SUPPLEMENTALRY MATERIAL

**Figure S1. THR-6E gene-level expression is concordantly elevated in SurvExpress-defined high-risk tumors.** Heatmap (**a**) and boxplots (**b**) showing expression of the six THR-6E genes across SurvExpress-defined low-risk (green) and high-risk (red) groups in the METABASE-10 breast cancer dataset. Risk grouping is derived from recurrence-free survival (RFS) using the SurvExpress risk model. Boxplots summarize gene-level expression in low-versus high-risk groups for each THR-6E component.

**Figure S2. THR-6E stratifies survival across breast cancer subgroups and shows limited prognostic signal in selected non-breast cancers.** Kaplan–Meier curves stratified by high versus low THR-6E mean expression using median cutoffs. Each plot reports hazard ratio (HR) with 95% confidence interval (CI) and two-sided log-rank p-value; numbers at risk are shown below curves.

(**a–b**) Overall survival (OS) and relapse-free survival (RFS) in ER+ /HER2– breast cancer, stratified by THR-6E expression.

(**c–d**) RFS in ER–/HER2– breast cancer and in HER2+ breast cancer, stratified by THR-6E expression.

(**e–f**) Progression-free survival (PFS) in ovarian cancer and OS in small cell carcinoma of the lung (SCLC), stratified by THR-6E expression.

**Figure S3. THR-6E expression in normal tissues is breast-enriched and maps to specific glandular epithelial cell subsets.**

**(a)** Tissue-level expression of ESR1 (ER), AR, and VDR and the six THR-6E gene products (KIF4A, KIF2C, CDC20, FAM64A/PIMREG, TPX2, and LMNB2) across normal human tissues, queried in ProteomicsDB.

**(b)** Cluster-level expression of THR-6E, ESR1 (ER), AR, VDR, and MKI67 (Ki67) across normal human breast glandular epithelial cell clusters (C1, C4, C9, C11, C16, C17, C20, C21) from the Human Protein Atlas (HPA) single-cell dataset (v23).

**Figure S4. Protein-level expression of THR-6E components in normal breast epithelium and breast cancer.** Representative immunohistochemistry (IHC) images from the Human Protein Atlas showing staining for proteins encoded by THR-6E genes in normal human breast tissue (**a**) and breast cancer tissue (**b**). Arrows indicate IHC-positive epithelial cells in normal breast.

**Figure S5. THR-6E gene–gene correlation structure is preserved from normal breast tissue to breast cancer.** Gene–gene correlation matrices for THR-6E, Oncotype DX, and PAM-50 gene sets computed from breast RNA-seq expression data in normal tissue (top panels) and breast tumors (bottom panels) using TNMplot. Colors represent Spearman correlation coefficients (−1 to 1).

**Figure S6. THR-6E genes are rarely altered in breast cancer compared with genes in commercial signatures.** Genomic alteration frequencies for THR-6E, Oncotype DX, and PAM-50 gene sets in breast cancer samples from TCGA-BRCA and METABRIC (total n=3,593), queried through cBioPortal. Alterations include mutations and copy-number changes as reported by the platform. THR-6E genes show low alteration frequency (mean 1.1% across the six genes), whereas Oncotype DX and PAM-50 include multiple genes with frequent copy-number gains in the range of 15–20% in queried genes.

**Figure S7. THR-6E is upregulated in primary and metastatic breast cancer relative to normal breast tissue.**

(a) Pan-cancer summary of THR-6E gene expression differences between tumors and matched normal tissues using RNA-seq data. Circles show log2 fold-change (tumor/normal); red indicates higher expression in tumors and blue indicates higher expression in normal tissues. Circle size is inversely proportional to the adjusted p-value.

(b) Breast-specific gene chip density plots showing log2 expression distributions of individual THR-6E genes across normal (red), primary tumor (green), and metastatic (blue) breast tissues.

(c) Breast-specific Targetgram plots (gene chip data) summarizing expression of individual THR-6E genes across normal, primary tumor, and metastatic breast tissues; segment size indicates the mean and dashed lines indicate the median within each tissue class.

(d) Violin plots (gene chip data) showing the combined THR-6E signature expression across normal, primary tumor, and metastatic breast tissues. Group differences were assessed by Kruskal–Wallis test (p=1.74×10⁻⁵²).

**Figure S8. THR-6E genes form a hormone-linked interaction network and connect to cancer-relevant phenotypes.**

(a) STRING protein association network for THR-6E components together with ESR1 (ER), AR, and VDR. Nodes represent proteins; colors distinguish query proteins from the first shell of interactors returned by STRING. Edges represent functional associations supported by curated databases, experimental evidence, and computational predictions (including gene neighborhood, gene fusion, co-occurrence, text mining, co-expression, and homology).

(b) CancerGeneNet (SIGNOR) regulatory network linking THR-6E genes to cancer-associated phenotypes. Query proteins are shown in yellow and first neighbors in green; protein families are shown as white circles and protein complexes as blue clover symbols. Solid edges denote direct interactions and dashed edges denote indirect interactions; blue arrows indicate up-regulation and red T-bars indicate down-regulation.

**Figure S9. THR-6E predicts chemotherapy response in grade III ER+/HER2– breast cancer, including node-positive disease.**

Top panels show THR-6E signature expression in chemotherapy responders versus non-responders among patients with grade III ER+/HER2– breast cancer. Bottom panels show receiver operating characteristic (ROC) curves evaluating discrimination performance, summarized by area under the ROC curve (AUC), as implemented by ROCplot. (**a**) All grade III ER+/HER2– breast cancers. (b) Grade III, ER+/HER2–/lymph node–positive (LN+) breast cancers.

**Figure S10. Cancer hallmark enrichment differs between THR-6E and Oncotype DX gene sets.**

Hallmark enrichment profiles for the indicated gene signatures computed relative to the platform-defined reference gene set. Each slice corresponds to one of the canonical cancer hallmarks; only hallmarks meeting statistical significance are colored. (**a**) THR-6E gene set. (**b**) Oncotype DX gene set. The accompanying table reports enriched pathways, gene overlap, adjusted p-values, and contributing genes.

## REFERENCES

1. Paik S, Shak S, Tang G, Kim C, Baker J, Cronin M, et al. A Multigene Assay to Predict Recurrence of Tamoxifen-Treated, Node-Negative Breast Cancer. New England Journal of Medicine. 2004 Dec 30;351(27):2817–26.

2. Paik S, Tang G, Shak S, Kim C, Baker J, Kim W, et al. Gene expression and benefit of chemotherapy in women with node-negative, estrogen receptor-positive breast cancer. J Clin Oncol. 2006 Aug 10;24(23):3726–34.

3. Albain KS, Barlow WE, Shak S, Hortobagyi GN, Livingston RB, Yeh IT, et al. Prognostic and predictive value of the 21-gene recurrence score assay in postmenopausal women with node-positive, oestrogen-receptor-positive breast cancer on chemotherapy: a retrospective analysis of a randomised trial. Lancet Oncol. 2010 Jan;11(1):55–65.

4. Sparano JA, Gray RJ, Makower DF, Pritchard KI, Albain KS, Hayes DF, et al. Prospective Validation of a 21-Gene Expression Assay in Breast Cancer. New England Journal of Medicine. 2015 Nov 19;373(21):2005–14.

5. Cardoso F, Veer LJ van’t, Bogaerts J, Slaets L, Viale G, Delaloge S, et al. 70-Gene Signature as an Aid to Treatment Decisions in Early-Stage Breast Cancer. New England Journal of Medicine. 2016 Aug 25;375(8):717–29.

6. Parker JS, Mullins M, Cheang MCU, Leung S, Voduc D, Vickery T, et al. Supervised Risk Predictor of Breast Cancer Based on Intrinsic Subtypes. J Clin Oncol. 2009 Mar 10;27(8):1160–7.

7. Emmert-Streib F. Grand Challenges for Artificial Intelligence in Molecular Medicine. Front Mol Med [Internet]. 2021 July 22 [cited 2026 Jan 28];1. Available from: https://www.frontiersin.org/journals/molecular-medicine/articles/10.3389/fmmed.2021.734659/full

8. Fertig EJ, Jaffee EM, Macklin P, Stearns V, Wang C. Forecasting cancer: from precision to predictive medicine. Med. 2021 Sept 10;2(9):1004–10.

9. Rogers TL, Johnson BJ, Munch SB. Chaos is not rare in natural ecosystems. Nat Ecol Evol. 2022 Aug;6(8):1105–11.

10. Torres R, Judson-Torres RL. Research Techniques Made Simple: Feature Selection for Biomarker Discovery. J Invest Dermatol. 2019 Oct;139(10):2068–2074.e1.

11. Omar M, Dinalankara W, Mulder L, Coady T, Zanettini C, Imada EL, et al. Using biological constraints to improve prediction in precision oncology. iScience. 2023 Mar 17;26(3):106108.

12. Kamran M, Bhattacharya U, Omar M, Marchionni L, Ince TA. ZNF92, an unexplored transcription factor with remarkably distinct breast cancer over-expression associated with prognosis and cell-of-origin. npj Breast Cancer. 2022 Aug 29;8(1):99.

13. Ince TA, Richardson AL, Bell GW, Saitoh M, Godar S, Karnoub AE, et al. Transformation of Different Human Breast Epithelial Cell Types Leads to Distinct Tumor Phenotypes. Cancer Cell. 2007 Aug 14;12(2):160–70.

14. Bhattacharya U, Kamran M, Manai M, Cristofanilli M, Ince TA. Cell-of-Origin Targeted Drug Repurposing for Triple-Negative and Inflammatory Breast Carcinoma with HDAC and HSP90 Inhibitors Combined with Niclosamide. Cancers (Basel). 2023 Jan 4;15(2):332.

15. Houseman EA, Ince TA. Normal cell-type epigenetics and breast cancer classification: a case study of cell mixture-adjusted analysis of DNA methylation data from tumors. Cancer Inform. 2014;13(Suppl 4):53–64.

16. Santagata S, Thakkar A, Ergonul A, Wang B, Woo T, Hu R, et al. Taxonomy of breast cancer based on normal cell phenotype predicts outcome. J Clin Invest. 2014 Feb 3;124(2):859–70.

17. Omar M, Harrell JC, Tamimi R, Marchionni L, Erdogan C, Nakshatri H, et al. A Triple Hormone Receptor ER, AR, and VDR Signature is a Robust Prognosis Predictor in Breast Cancer. Breast Cancer Research. 2024 Aug;

18. Szklarczyk D, Kirsch R, Koutrouli M, Nastou K, Mehryary F, Hachilif R, et al. The STRING database in 2023: protein–protein association networks and functional enrichment analyses for any sequenced genome of interest. Nucleic Acids Research. 2023 Jan 6;51(D1):D638–46.

19. Cao Q, Song Z, Ruan H, Wang C, Yang X, Bao L, et al. Targeting the KIF4A/AR Axis to Reverse Endocrine Therapy Resistance in Castration-resistant Prostate Cancer. Clin Cancer Res. 2020 Mar 15;26(6):1516–28.

20. Liu S, Ye Z, Xue VW, Sun Q, Li H, Lu D. KIF2C is a prognostic biomarker associated with immune cell infiltration in breast cancer. BMC Cancer. 2023 Apr 4;23(1):307.

21. Alfarsi LH, Ansari RE, Craze ML, Toss MS, Masisi B, Ellis IO, et al. CDC20 expression in oestrogen receptor positive breast cancer predicts poor prognosis and lack of response to endocrine therapy. Breast Cancer Res Treat. 2019 Dec;178(3):535–44.

22. Zhou Y, Ou L, Xu J, Yuan H, Luo J, Shi B, et al. FAM64A is an androgen receptor-regulated feedback tumor promoter in prostate cancer. Cell Death Dis. 2021 July 2;12(7):668.

23. Sun B, Long Y, Xiao L, Wang J, Yi Q, Tong D, et al. Target Protein for Xklp2 Functions as Coactivator of Androgen Receptor and Promotes the Proliferation of Prostate Carcinoma Cells. J Oncol. 2022;2022:6085948.

24. Novaro V, Roskelley CD, Bissell MJ. Collagen-IV and laminin-1 regulate estrogen receptor alpha expression and function in mouse mammary epithelial cells. J Cell Sci. 2003 July 15;116(Pt 14):2975–86.

25. Curtis C, Shah SP, Chin SF, Turashvili G, Rueda OM, Dunning MJ, et al. The genomic and transcriptomic architecture of 2,000 breast tumours reveals novel subgroups. Nature. 2012 Apr 18;486(7403):346–52.

26. Schmidt T, Samaras P, Frejno M, Gessulat S, Barnert M, Kienegger H, et al. ProteomicsDB. Nucleic Acids Res. 2018 Jan 4;46(D1):D1271–81.

27. Karlsson M, Zhang C, Méar L, Zhong W, Digre A, Katona B, et al. A single–cell type transcriptomics map of human tissues. Science Advances. 2021 July 28;7(31):eabh2169.

28. Uhlén M, Fagerberg L, Hallström BM, Lindskog C, Oksvold P, Mardinoglu A, et al. Tissue-based map of the human proteome. Science. 2015 Jan 23;347(6220):1260419.

29. Bartha Á, Győrffy B. TNMplot.com: A Web Tool for the Comparison of Gene Expression in Normal, Tumor and Metastatic Tissues. Int J Mol Sci. 2021 Mar 5;22(5):2622.

30. Zou JX, Duan Z, Wang J, Sokolov A, Xu J, Chen CZ, et al. Kinesin family deregulation coordinated by bromodomain protein ANCCA and histone methyltransferase MLL for breast cancer cell growth, survival, and tamoxifen resistance. Mol Cancer Res. 2014 Apr;12(4):539–49.

31. Iannuccelli M, Micarelli E, Surdo PL, Palma A, Perfetto L, Rozzo I, et al. CancerGeneNet: linking driver genes to cancer hallmarks. Nucleic Acids Res. 2020 Jan 8;48(D1):D416–21.

32. Andre F, Ismaila N, Allison KH, Barlow WE, Collyar DE, Damodaran S, et al. Biomarkers for Adjuvant Endocrine and Chemotherapy in Early-Stage Breast Cancer: ASCO Guideline Update. J Clin Oncol. 2022 June;40(16):1816–37.

33. Wang H, Yee D. I-SPY 2: a Neoadjuvant Adaptive Clinical Trial Designed to Improve Outcomes in High-Risk Breast Cancer. Current breast cancer reports. 2019 Nov 20;11(4):303.

34. I-SPY2 Trial Consortium. Association of Event-Free and Distant Recurrence–Free Survival With Individual-Level Pathologic Complete Response in Neoadjuvant Treatment of Stages 2 and 3 Breast Cancer: Three-Year Follow-up Analysis for the I-SPY2 Adaptively Randomized Clinical Trial. JAMA Oncol. 2020 Sept 1;6(9):1355–62.

35. Smith JC, Sheltzer JM. Genome-wide identification and analysis of prognostic features in human cancers. Cell Rep. 2022 Mar 29;38(13):110569.

36. Hoadley KA, Yau C, Hinoue T, Wolf DM, Lazar AJ, Drill E, et al. Cell-of-Origin Patterns Dominate the Molecular Classification of 10,000 Tumors from 33 Types of Cancer. Cell. 2018 Apr 5;173(2):291–304.e6.

37. Polak P, Karlić R, Koren A, Thurman R, Sandstrom R, Lawrence MS, et al. Cell-of-origin chromatin organization shapes the mutational landscape of cancer. Nature. 2015 Feb;518(7539):360–4.

38. Houseman EA, Kile ML, Christiani DC, Ince TA, Kelsey KT, Marsit CJ. Reference-free deconvolution of DNA methylation data and mediation by cell composition effects. BMC Bioinformatics. 2016 June 29;17(1):259.

39. Kalinsky K, Barlow WE, Gralow JR, Meric-Bernstam F, Albain KS, Hayes DF, et al. 21-Gene Assay to Inform Chemotherapy Benefit in Node-Positive Breast Cancer. New England Journal of Medicine. 2021 Dec 15;385(25):2336–47.

40. Venet D, Dumont JE, Detours V. Most Random Gene Expression Signatures Are Significantly Associated with Breast Cancer Outcome. PLOS Computational Biology. 2011 Oct 20;7(10):e1002240.

41. Hanahan D, Weinberg RA. Hallmarks of cancer: the next generation. Cell. 2011 Mar 4;144(5):646–74.

42. Hanahan D. Hallmarks of Cancer: New Dimensions. Cancer Discov. 2022 Jan 12;12(1):31–46.

43. Lánczky A, Győrffy B. Web-Based Survival Analysis Tool Tailored for Medical Research (KMplot): Development and Implementation. J Med Internet Res. 2021 July 26;23(7):e27633.

44. Fekete JT, Győrffy B. ROCplot.org: Validating predictive biomarkers of chemotherapy/hormonal therapy/anti-HER2 therapy using transcriptomic data of 3,104 breast cancer patients. International Journal of Cancer. 2019;145(11):3140–51.

45. Aguirre-Gamboa R, Gomez-Rueda H, Martínez-Ledesma E, Martínez-Torteya A, Chacolla-Huaringa R, Rodriguez-Barrientos A, et al. SurvExpress: an online biomarker validation tool and database for cancer gene expression data using survival analysis. PLoS One. 2013;8(9):e74250.

46. Bhat-Nakshatri P, Gao H, Sheng L, McGuire PC, Xuei X, Wan J, et al. A single-cell atlas of the healthy breast tissues reveals clinically relevant clusters of breast epithelial cells. Cell Rep Med. 2021 Mar 16;2(3):100219.

