## Supplementary figures and images for "THR-6E: A Six-Gene Cell-of-Origin Signature Stratifies Risk and Predicts Systemic Therapy Response in ER+/HER2− Breast Cancer"

### Figure S1

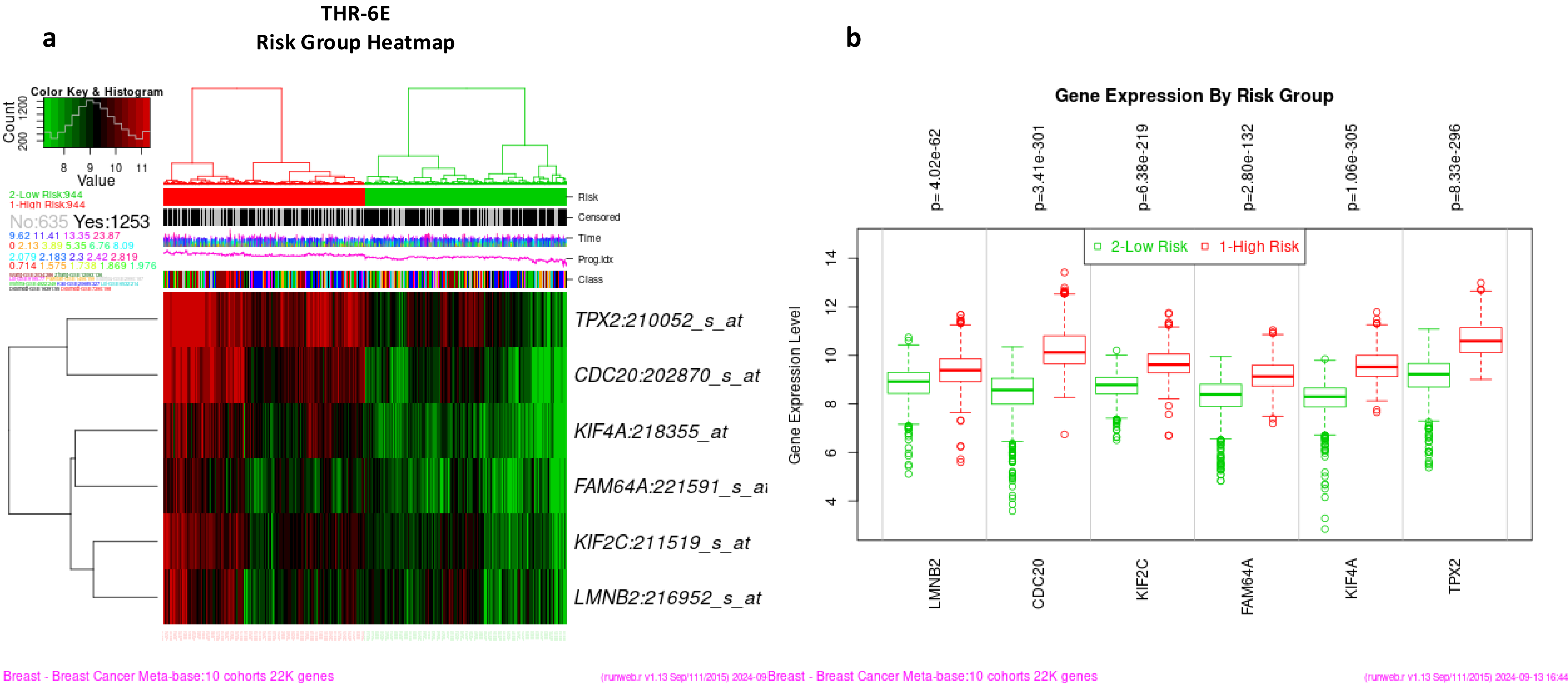

### Figure S2

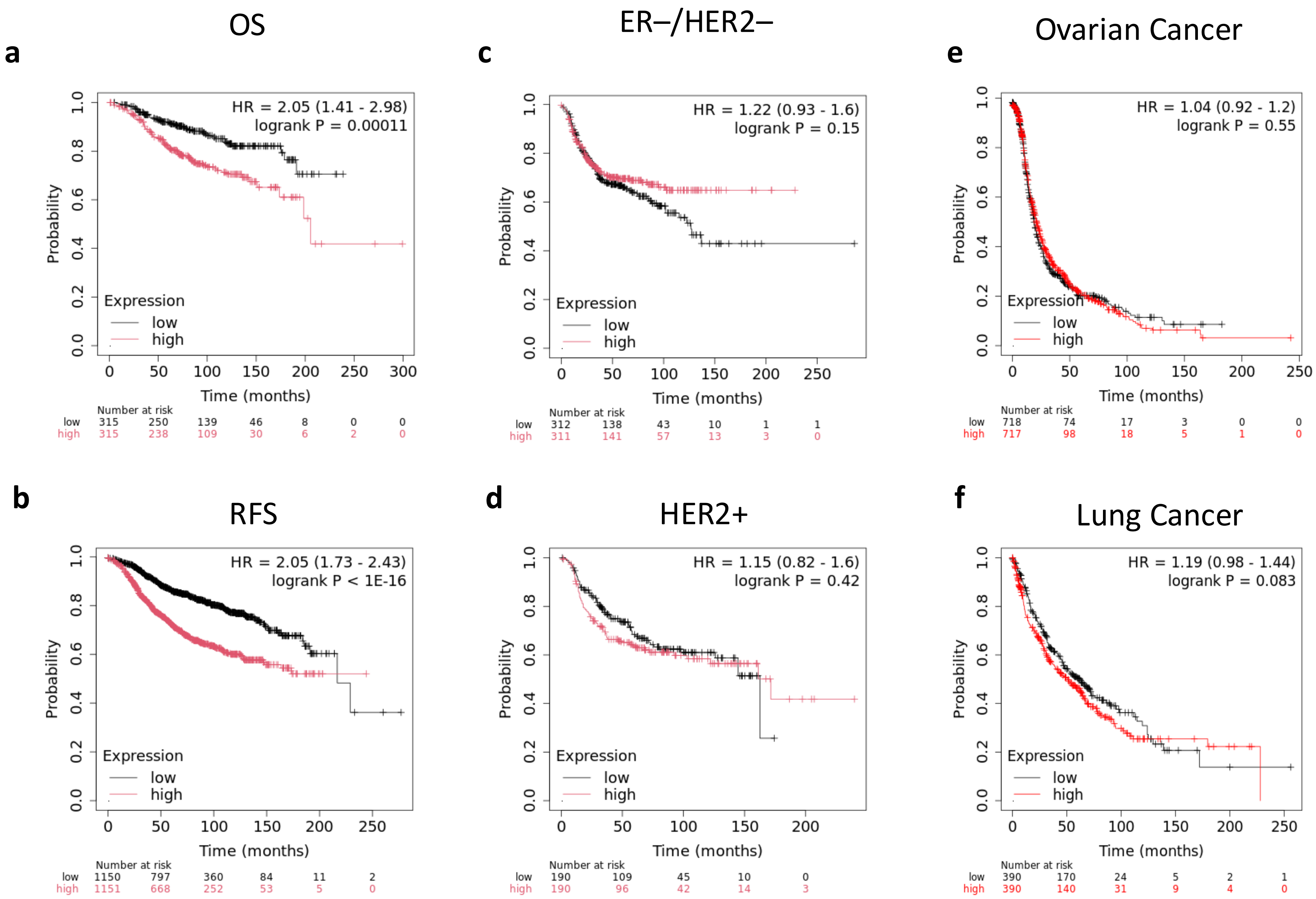

### Figure S3

Fig. S3

**a**

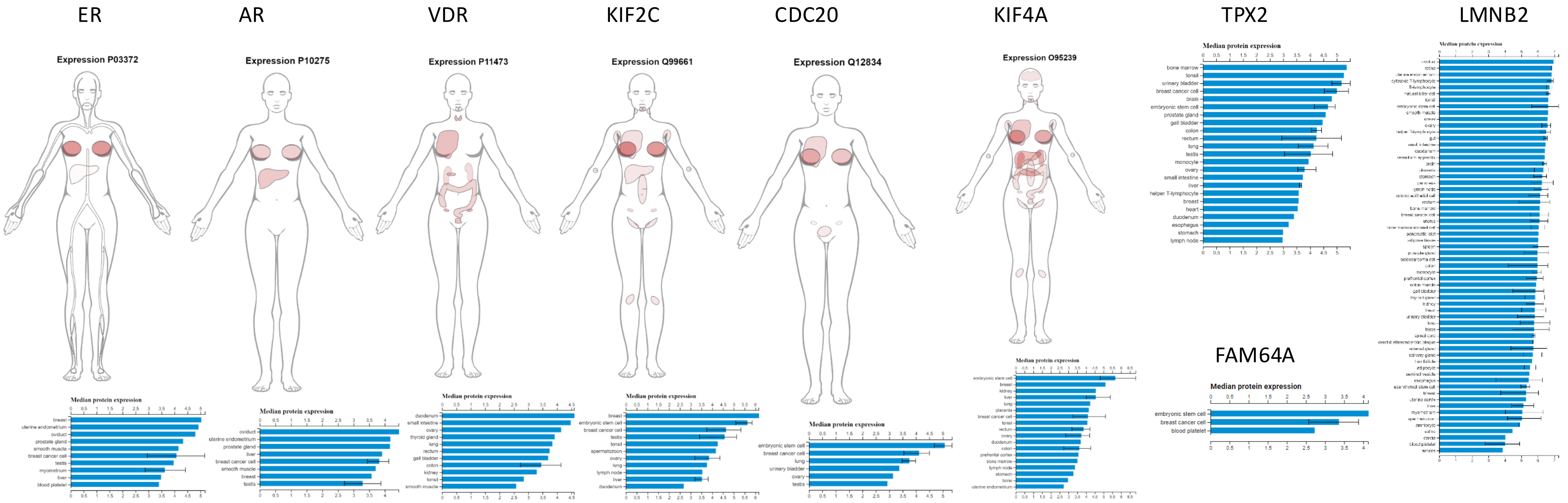


**b**

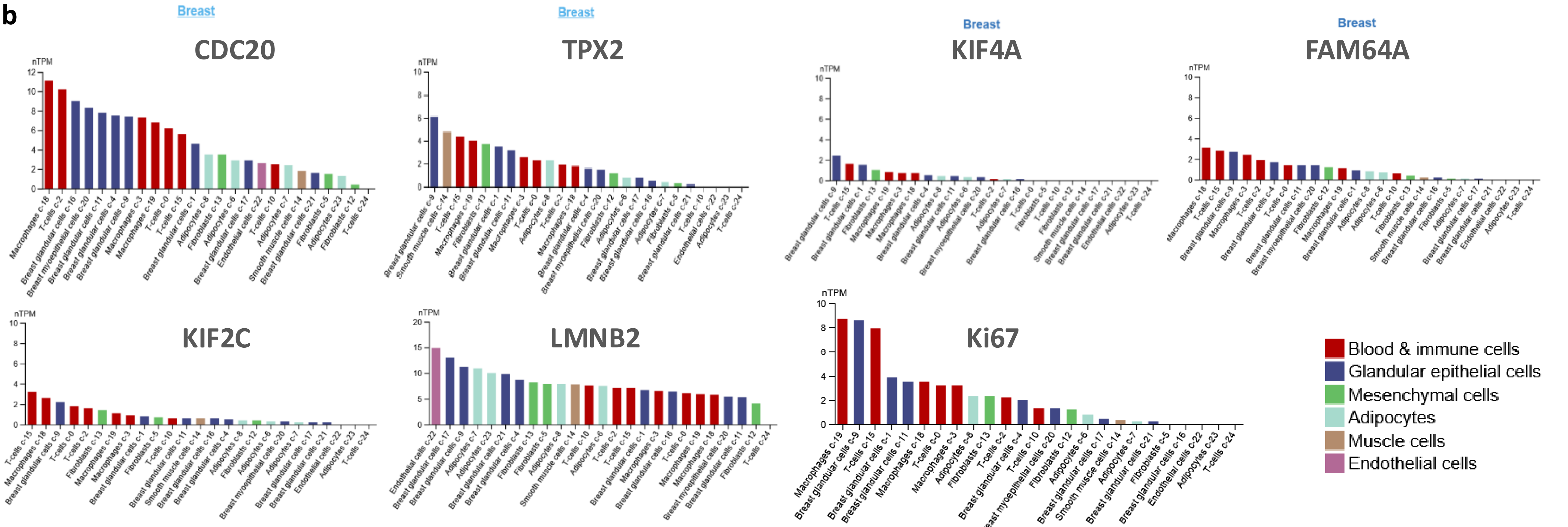


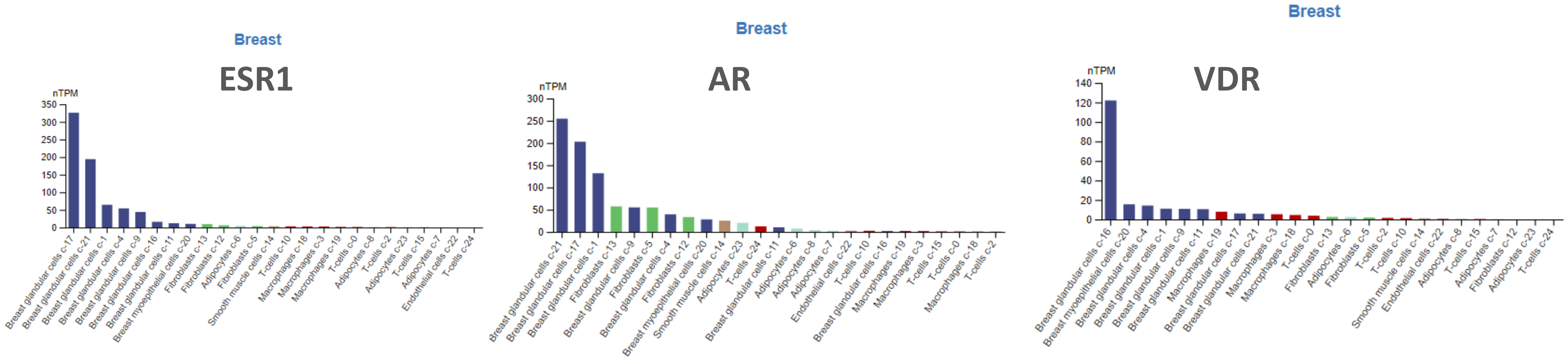


| Cluster | THR-6E | | | | | | |
|---|---|---|---|---|---|---|---|
| | CDC20 | FAM64A | KIF2C | KIF4A | LMNB2 | TPX2 | Total |
| c-9 | √ | √ | √ | √ | √ | √ | 6 |
| c-4 | √ | √ | | | √ | | 3 |
| c-1 | √ | | | √ | | | 2 |
| c-11 | √ | √ | | | | | 2 |
| c-20 | √ | √ | | | | | 2 |
| c-16 | √ | | | | | | 1 |
| c-17 | | | | | √ | | 1 |
| c-21 | | | | | √ | | 1 |

### Figure S4

Fig. S4

**a**

##### Normal Human Breast

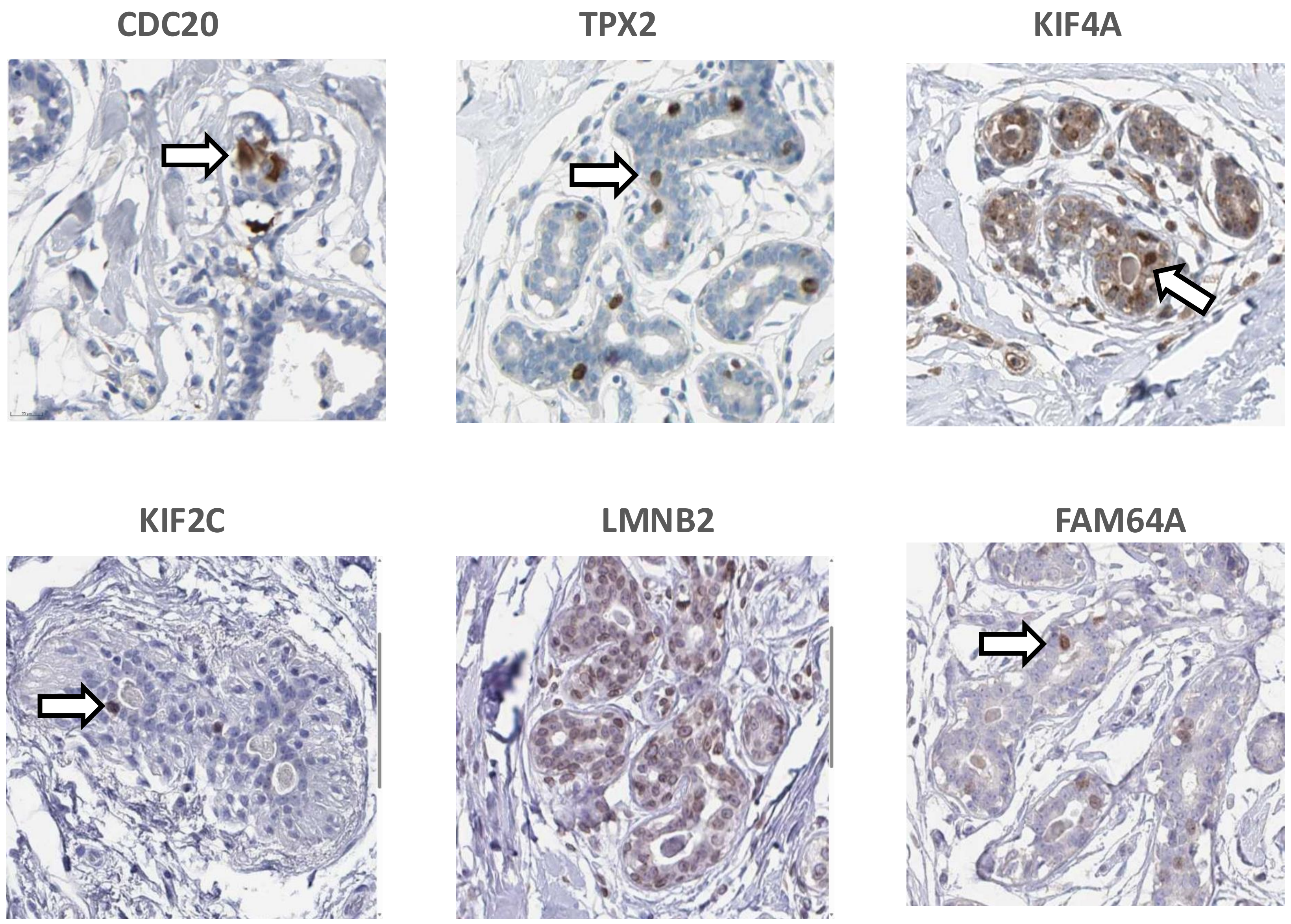


**b**

##### Breast Cancer

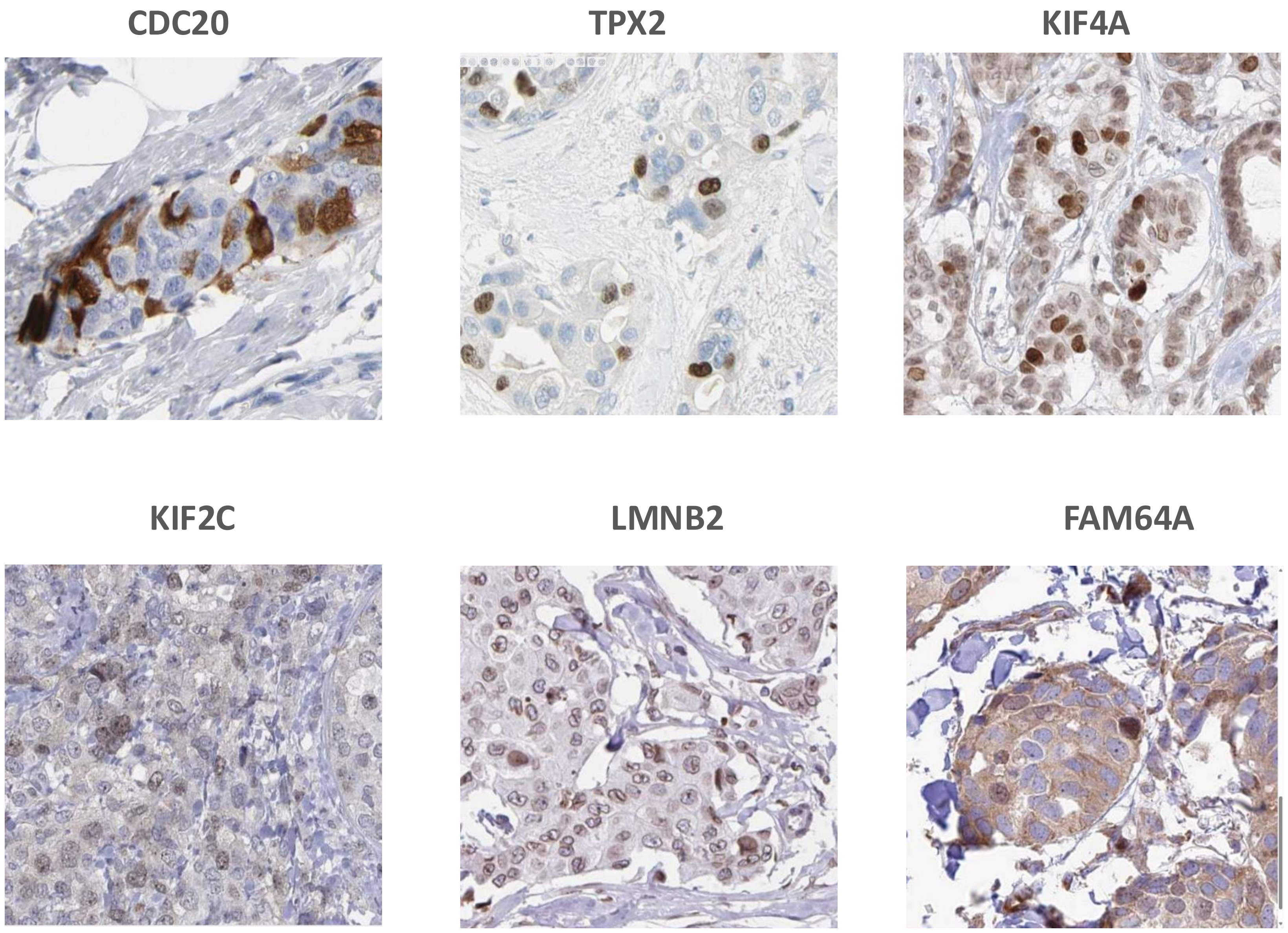

### Figure S5

Fig. S5

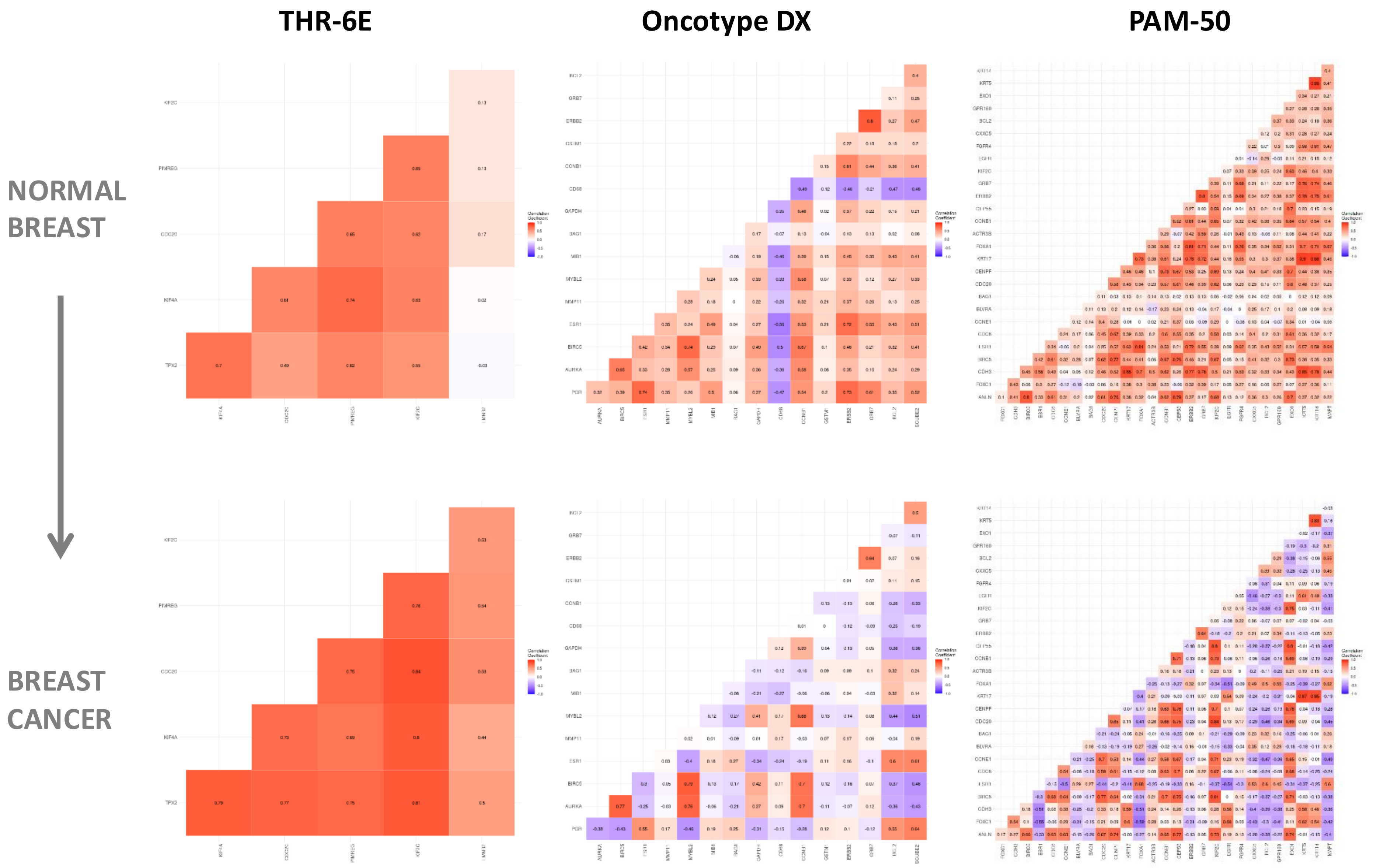

### Figure S6

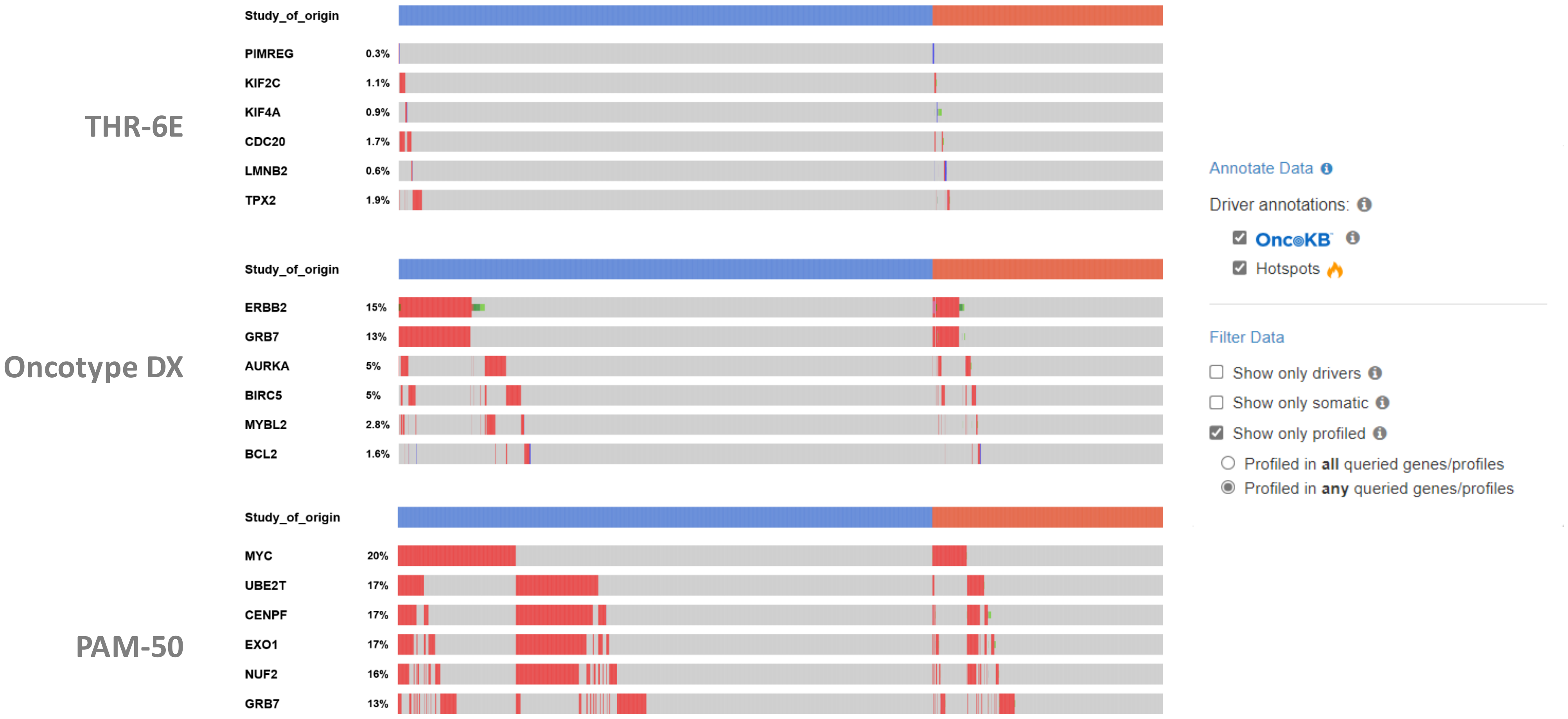

### Figure S7

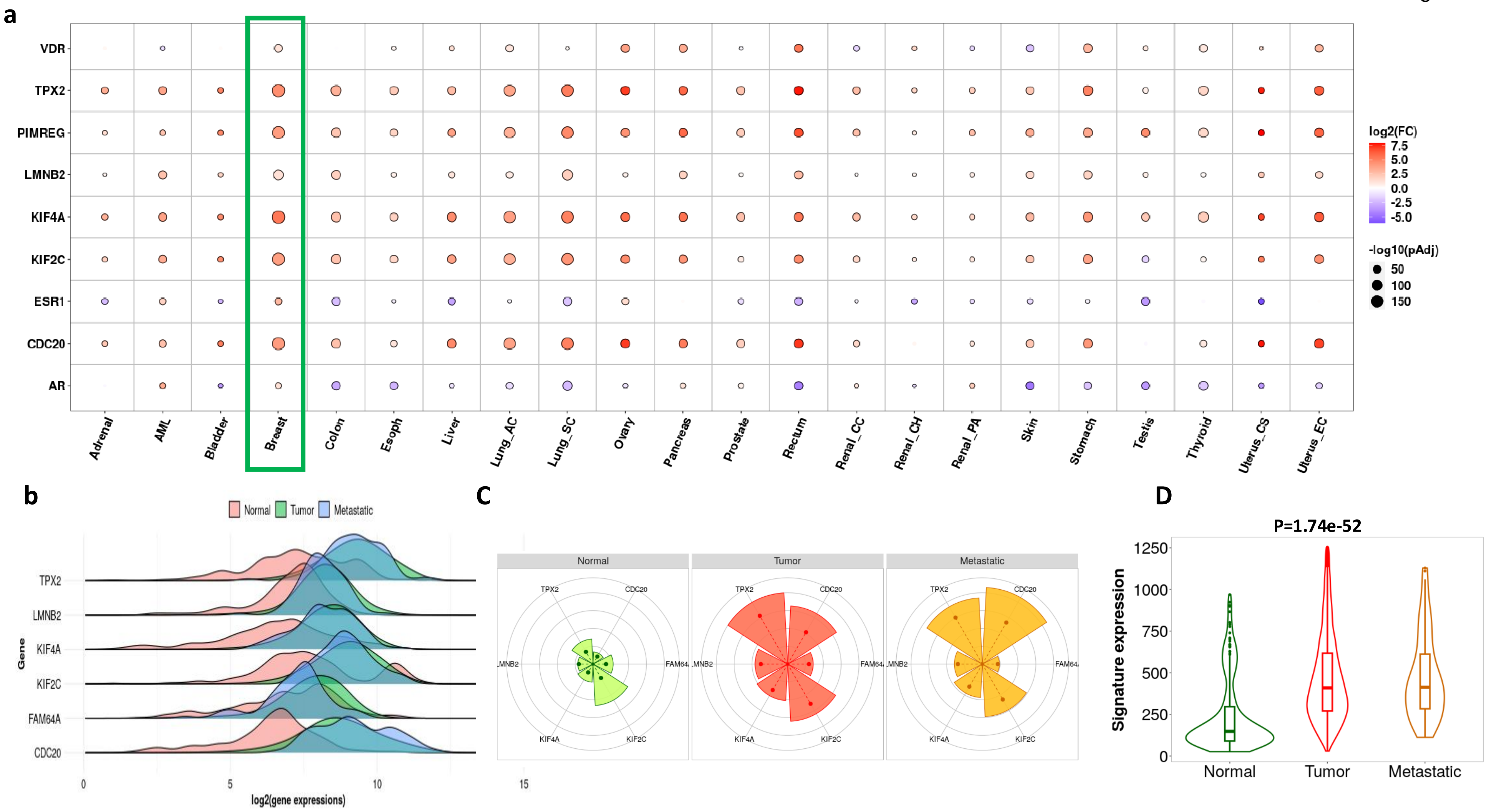

### Figure S8

#### THR-6E Interactome

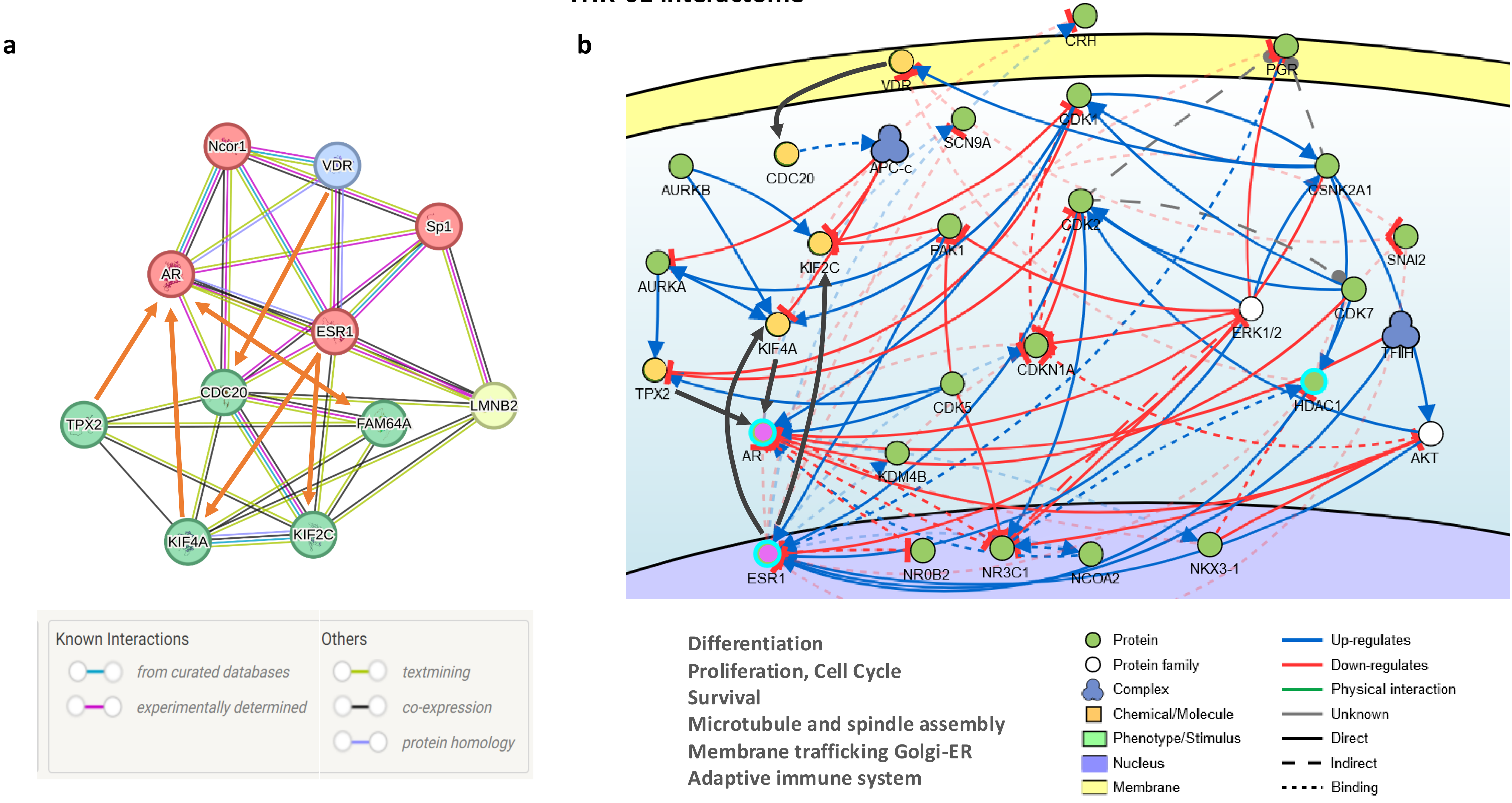
