## Supplementary material for "THR-6E: A Six-Gene Cell-of-Origin Signature Stratifies Risk and Predicts Systemic Therapy Response in ER+/HER2− Breast Cancer": Figure S9

### a Grade III Chemotherapy: AUC=0.622

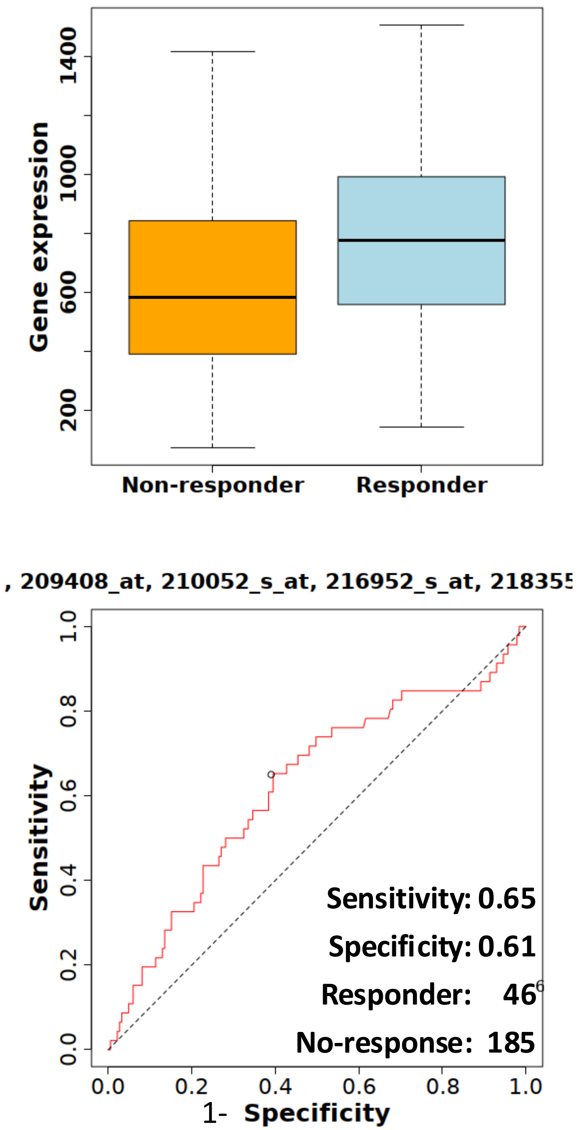


### b Grade III LN + Chemotherapy: AUC =0.667

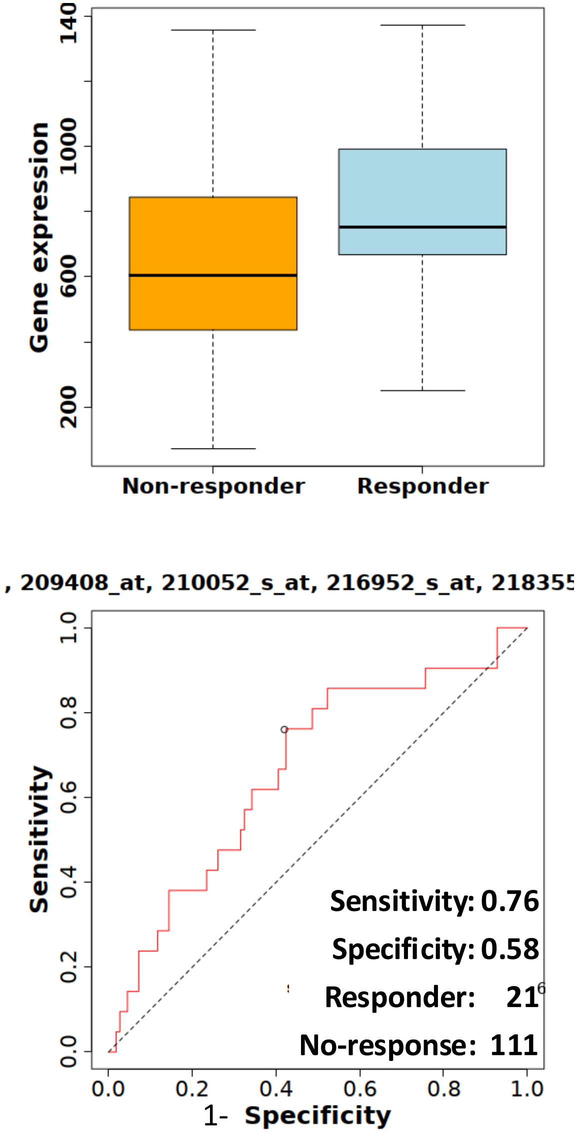
