## Supplementary material for "THR-6E: A Six-Gene Cell-of-Origin Signature Stratifies Risk and Predicts Systemic Therapy Response in ER+/HER2− Breast Cancer": Figure S10

Fig. S10

### Cancer Hallmarks Enrichment

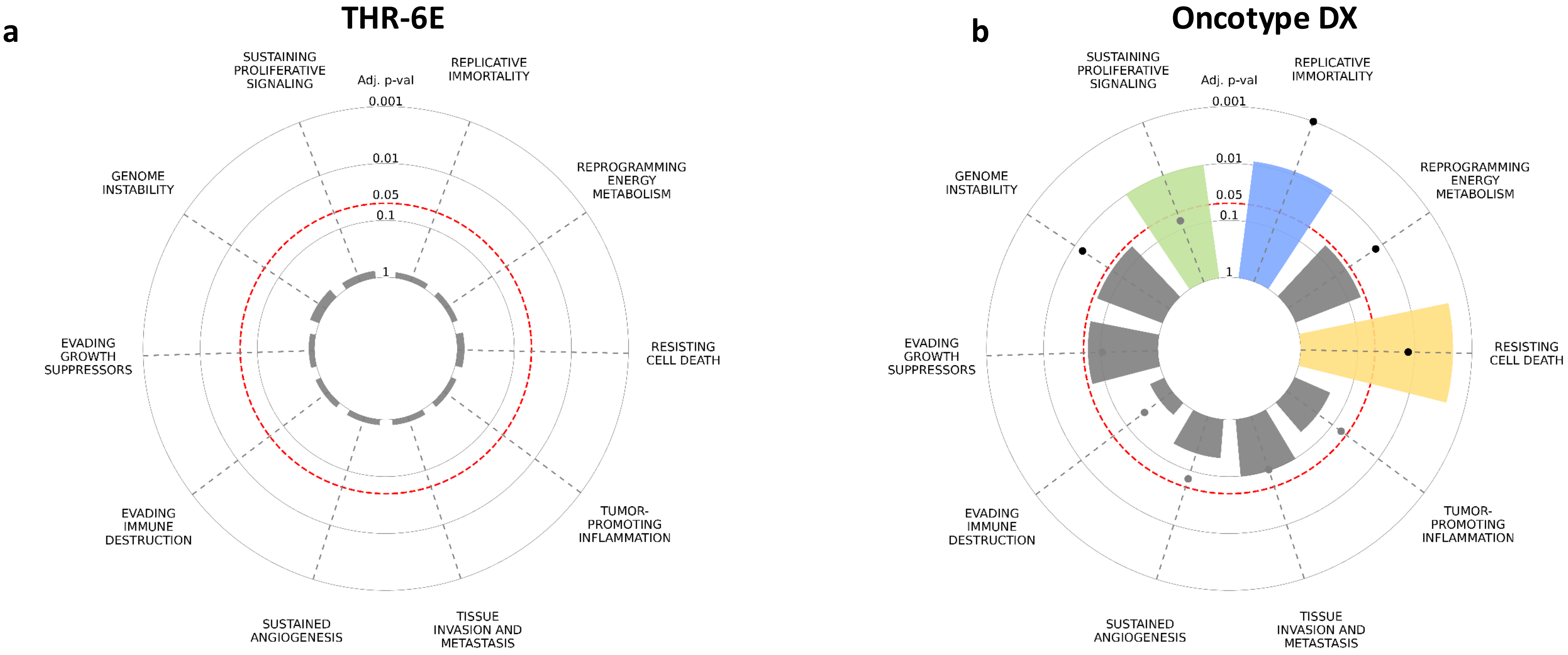


| | THR-6E | | | Oncotype | | |
|---|---|---|---|---|---|---|
| Term | Overlap | Adjusted P-value | Genes | Overlap | Adjusted P-value | Genes |
| **SUSTAINING PROLIFERATIVE SIGNALING** | 1 | 0.74 | CDC20 | **9** | **0.010** | **AURKA;CCNB1;GRB7;PGR;BIRC5;BCL2;ESR1;ERBB2;SCUBE2** |
| **RESISTING CELL DEATH** | 1 | 0.74 | LMNB2 | **8** | **0.002** | **AURKA;CCNB1;CD68;BAG1;BIRC5;BCL2;ESR1;ERBB2** |
| **REPLICATIVE IMMORTALITY** | 0 | 1.00 | | **4** | **0.009** | **BCL2;ESR1;CCNB1;ERBB2** |
| EVADING GROWTH SUPPRESSORS | 0 | 1.00 | | 7 | 0.060 | GRB7;PGR;BIRC5;BCL2;ESR1;ERBB2;SCUBE2 |
| TISSUE INVASION AND METASTASIS | 0 | 1.00 | | 5 | 0.098 | GRB7;BCL2;ESR1;ERBB2;MMP11 |
| GENOME INSTABILITY | 1 | 0.68 | CDC20 | 3 | 0.060 | ESR1;CCNB1;GSTM1 |
| EVADING IMMUNE DESTRUCTION | 0 | 1.00 | | 1 | 0.536 | ESR1 |
| SUSTAINED ANGIOGENESIS | 0 | 1.00 | | 2 | 0.208 | ERBB2;ESR1 |
| TUMOR-PROMOTING INFLAMMATION | 0 | 1.00 | | 2 | 0.208 | BCL2;ESR1 |
| REPROGRAMMING ENERGY METABOLISM | 0 | 1.00 | | 3 | 0.060 | BCL2;CCNB1;ERBB2 |
